# Sensitive Pathogen Detection and Drug Resistance Characterization Using Pathogen-Derived Enzyme Activity Amplified by LAMP or CRISPR-Cas

**DOI:** 10.1101/2024.03.29.24305085

**Authors:** Qin Wang, Enos C. Kline, Shane D. Gilligan-Steinberg, James J. Lai, Ian T. Hull, Ayokunle O. Olanrewaju, Nuttada Panpradist, Barry R. Lutz

## Abstract

Pathogens encapsulate or encode their own suite of enzymes to facilitate replication in the host. The pathogen-derived enzymes possess specialized activities that are essential for pathogen replication and have naturally been candidates for drug targets. Phenotypic assays detecting the activities of pathogen-derived enzymes and characterizing their inhibition under drugs offer an opportunity for pathogen detection, drug resistance testing for individual patients, and as a research tool for new drug development. Here, we used HIV as an example to develop assays targeting the reverse transcriptase (RT) enzyme encapsulated in HIV for sensitive detection and phenotypic characterization, with the potential for point-of-care (POC) applications. Specifically, we targeted the complementary (cDNA) generation activity of the HIV RT enzyme by adding engineered RNA as substrates for HIV RT enzyme to generate cDNA products, followed by cDNA amplification and detection facilitated by loop-mediated isothermal amplification (LAMP) or CRISPR-Cas systems. To guide the assay design, we first used qPCR to characterize the cDNA generation activity of HIV RT enzyme. In the LAMP-mediated Product-Amplified RT activity assay (LamPART), the cDNA generation and LAMP amplification were combined into one pot with novel assay designs. When coupled with direct immunocapture of HIV RT enzyme for sample preparation and endpoint lateral flow assays for detection, LamPART detected as few as 20 copies of HIV RT enzyme spiked into 25μL plasma (fingerstick volume), equivalent to a single virion. In the Cas-mediated Product-Amplified RT activity assay (CasPART), we tailored the substrate design to achieve a LoD of 2e4 copies (1.67fM) of HIV RT enzyme. Furthermore, with its phenotypic characterization capability, CasPART was used to characterize the inhibition of HIV RT enzyme under antiretroviral drugs and differentiate between wild-type and mutant HIV RT enzyme for potential phenotypic drug resistance testing. Moreover, the CasPART assay can be readily adapted to target the activity of other pathogen-derived enzymes. As a proof-of-concept, we successfully adapted CasPART to detect HIV integrase with a sensitivity of 83nM. We anticipate the developed approach of detecting enzyme activity with product amplification has the potential for a wide range of pathogen detection and phenotypic characterization.

## Introduction

Replication of pathogens in the host relies on their own suite of enzymes, either encapsulated within the pathogen (e.g., bacteria [1], HIV [2]) or encoded in their genome and subsequently synthesized in host cells (e.g., SARS-CoV-2 [3], HCV [4]). The pathogen-derived enzymes have evolved to facilitate critical replication steps and thereby possess specialized activities, such as reverse transcription, DNA or RNA polymerization, and DNA topology adjustment (Table 1). Being specific to the pathogens and critical in their replication, the enzyme activities are naturally candidates for drug targets [5,6]. The ongoing challenges of both antiviral and antimicrobial resistance highlight the pressing need for pathogen drug resistance testing [5,7]. Detecting the activities of pathogen-derived enzymes offers an opportunity for pathogen detection, and measuring their activities in the presence of drugs provides direct characterization of drug resistance. Here, we explore developing pathogen-derived enzyme activity assays, where engineered nucleic acids are added as enzyme substrates and the enzyme products are amplified with molecular assays, for sensitive pathogen detection and drug resistance characterization. In pathogen-derived enzyme activity assays, the product amplification through molecular assays provides a higher sensitivity compared to antigen or antibody immunoassays, and the engineered substrate sequence bypasses the challenges in pathogen nucleic acid testing posed by endogenous sequence restrictions and genetic mutations. Moreover, the pathogen-derived enzyme activity assay with spiked drugs could present a direct measure of drug efficacy, which is valuable to treatment selection for individual patients and research development for new drugs.

**Table 1.**
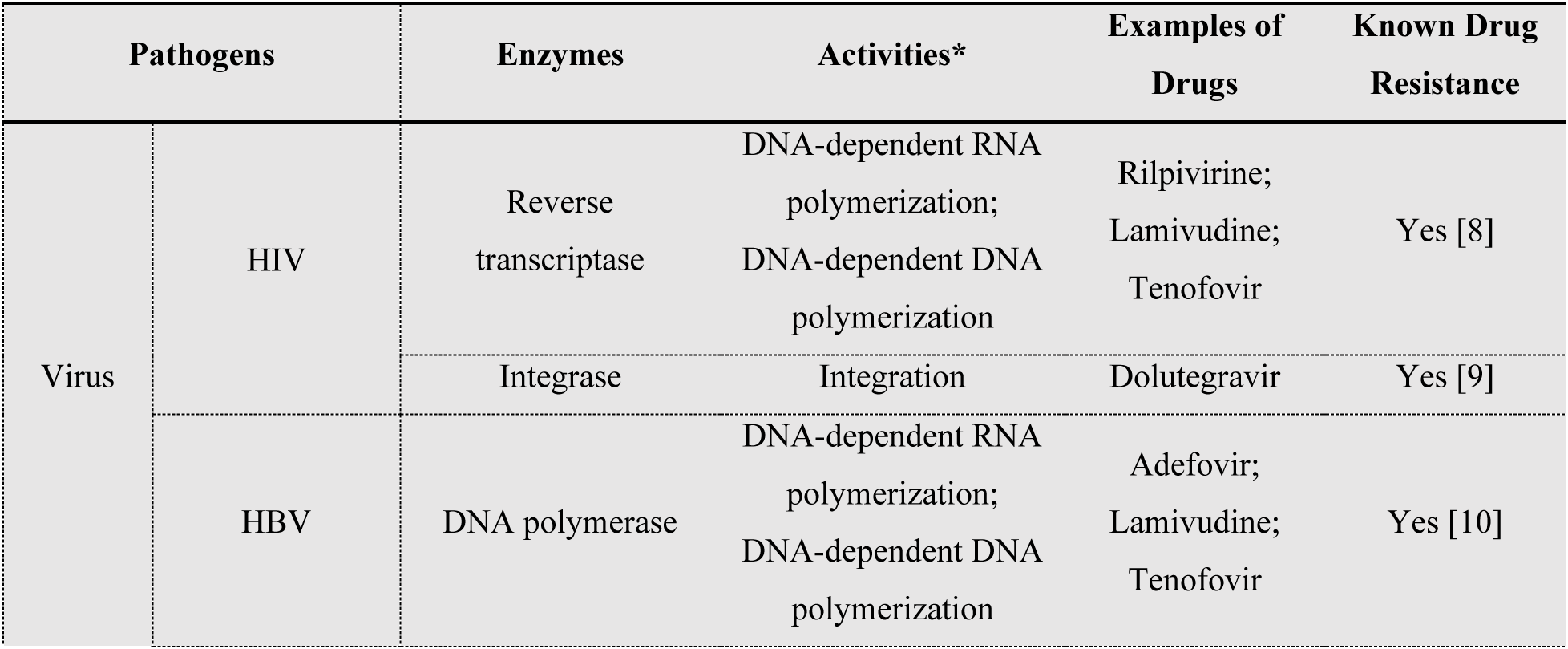
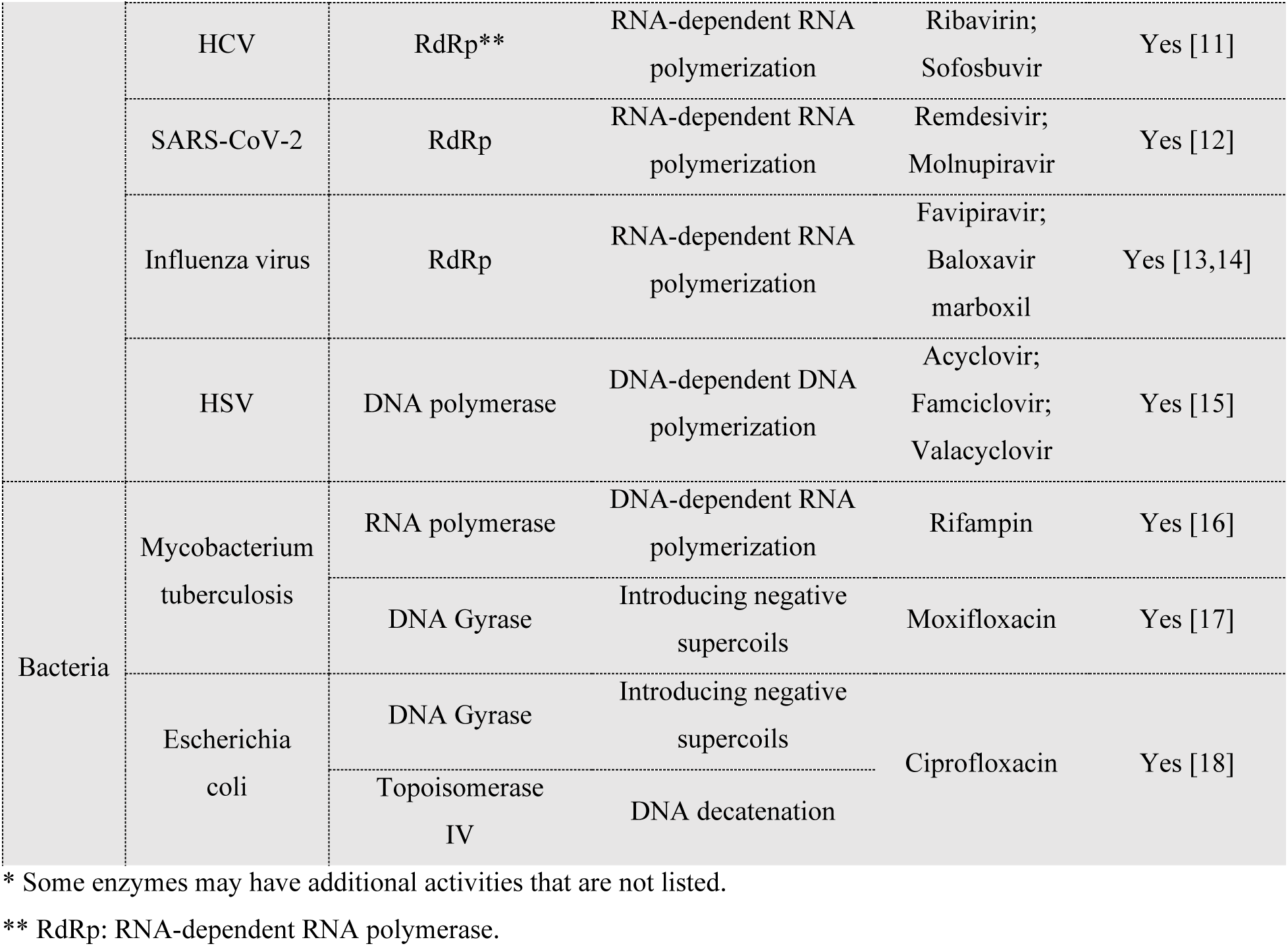
Examples of pathogen-derived enzymes and their activities, drugs, and known resistance.

| Pathogens |  | Enzymes | Activities* | Examples of Drugs | Known Drug Resistance |
| --- | --- | --- | --- | --- | --- |
| Virus | HIV | Reverse transcriptase | DNA-dependent RNA polymerization;<br>DNA-dependent DNA polymerization | Rilpivirine;<br>Lamivudine;<br>Tenofovir | Yes [8] |
|  |  | Integrase | Integration | Dolutegravir | Yes [9] |
|  | HBV | DNA polymerase | DNA-dependent RNA polymerization;<br>DNA-dependent DNA polymerization | Adefovir;<br>Lamivudine;<br>Tenofovir | Yes [10] |
|  | HCV | RdRp** | RNA-dependent RNA polymerization | Ribavirin;<br>Sofosbuvir | Yes [11] |
|  | SARS-CoV-2 | RdRp | RNA-dependent RNA polymerization | Remdesivir;<br>Molnupiravir | Yes [12] |
|  | Influenza virus | RdRp | RNA-dependent RNA polymerization | Favipiravir;<br>Baloxavir<br>marboxil | Yes [13,14] |
|  | HSV | DNA polymerase | DNA-dependent DNA polymerization | Acyclovir;<br>Famciclovir;<br>Valacyclovir | Yes [15] |
| Bacteria | Mycobacterium tuberculosis | RNA polymerase | DNA-dependent RNA polymerization | Rifampin | Yes [16] |
|  |  | DNA Gyrase | Introducing negative supercoils | Moxifloxacin | Yes [17] |
|  | Escherichia coli | DNA Gyrase | Introducing negative supercoils | Ciprofloxacin | Yes [18] |
|  |  | Topoisomerase IV | DNA decatenation |  |  |

\* Some enzymes may have additional activities that are not listed.
\*\* RdRp: RNA-dependent RNA polymerase.

We used HIV as an example due to the persistent need for sensitive HIV detection and drug resistance testing at the point of care (POC), especially in low-resource settings [19]. Currently, the HIV RNA reverse transcription-polymerase chain reaction (rt-PCR) assay is the most sensitive method for HIV detection and the first step in genotypic HIV drug resistance tests [20,21]. However, HIV RNA as the target biomarker presents significant challenges for POC applications, including extremely low copy number in samples with low viral load, susceptibility to highly abundant plasma ribonucleases (RNase), resource-intensive extraction processes, and sequence diversity among subtypes resulting in varied assay performance [22–24]. HIV, as a retrovirus, carries reverse transcriptase (RT) enzyme that possesses RNA-dependent DNA polymerase activity to convert its RNA genome into complementary DNA (cDNA) [25]. Multiple assays have been developed in the past to detect the cDNA generation activity of HIV RT enzyme for sensitive HIV detection, and the assays were further applied to viral load monitoring and drug resistance testing [26–31]. Compared to HIV RNA, the HIV RT enzyme offers several advantages as a biomarker for the POC. First, each HIV virion is reported to have 50-100 copies of RT enzyme, in contrast to two copies of HIV RNA, providing more targets available for detection [25,32]. In addition,

HIV RT enzyme as a protein is inert to RNases in plasma and could potentially be purified by immunocapture, facilitating simpler sample preparation. Moreover, the conserved RT enzyme activity among HIV subtypes allows a broad coverage of HIV without complication from diverse genetic sequences. Lastly, HIV RT enzyme is a primary target for antiretroviral therapy (ART). Assessing the activity of HIV RT enzyme under ART drugs can be used for phenotypic drug resistance testing and provide a direct measure of the ART efficacy without genetic interpretations [33]. However, previous RT activity assays used high-resource methods such as ultracentrifugation to purify the virus from samples and PCR to amplify and detect the cDNA generated by HIV RT enzyme, thus limiting their use to higher-tier laboratories. In addition, the assays involved multiple separate steps including cDNA generation, RNA substrate digestion, and PCR amplification, adding to assay complication and hindering POC use.

Loop-mediated isothermal amplification (LAMP) is a highly sensitive approach for rapid DNA amplification at a constant temperature [34]. In contrast to PCR, LAMP only requires a single-temperature heater without thermal cycling, making it suitable for the POC [35]. Here, we developed a LAMP-mediated Product-Amplified Reverse Transcriptase activity assay (LamPART) to amplify and detect cDNA generated by HIV RT enzyme for simple yet sensitive HIV detection (Schematic 1). While previous HIV RT activity assays needed to separate the cDNA generation and amplification steps, here we combined them into one pot with novel assay designs to simplify the workflow. Moreover, we developed direct immunocapture of HIV RT enzyme for sample preparation and endpoint lateral flow assays for detection. The integrated sample-to-result workflow was further validated using HIV-negative plasma spiked with HIV RT enzyme.

**Schematic 1.**
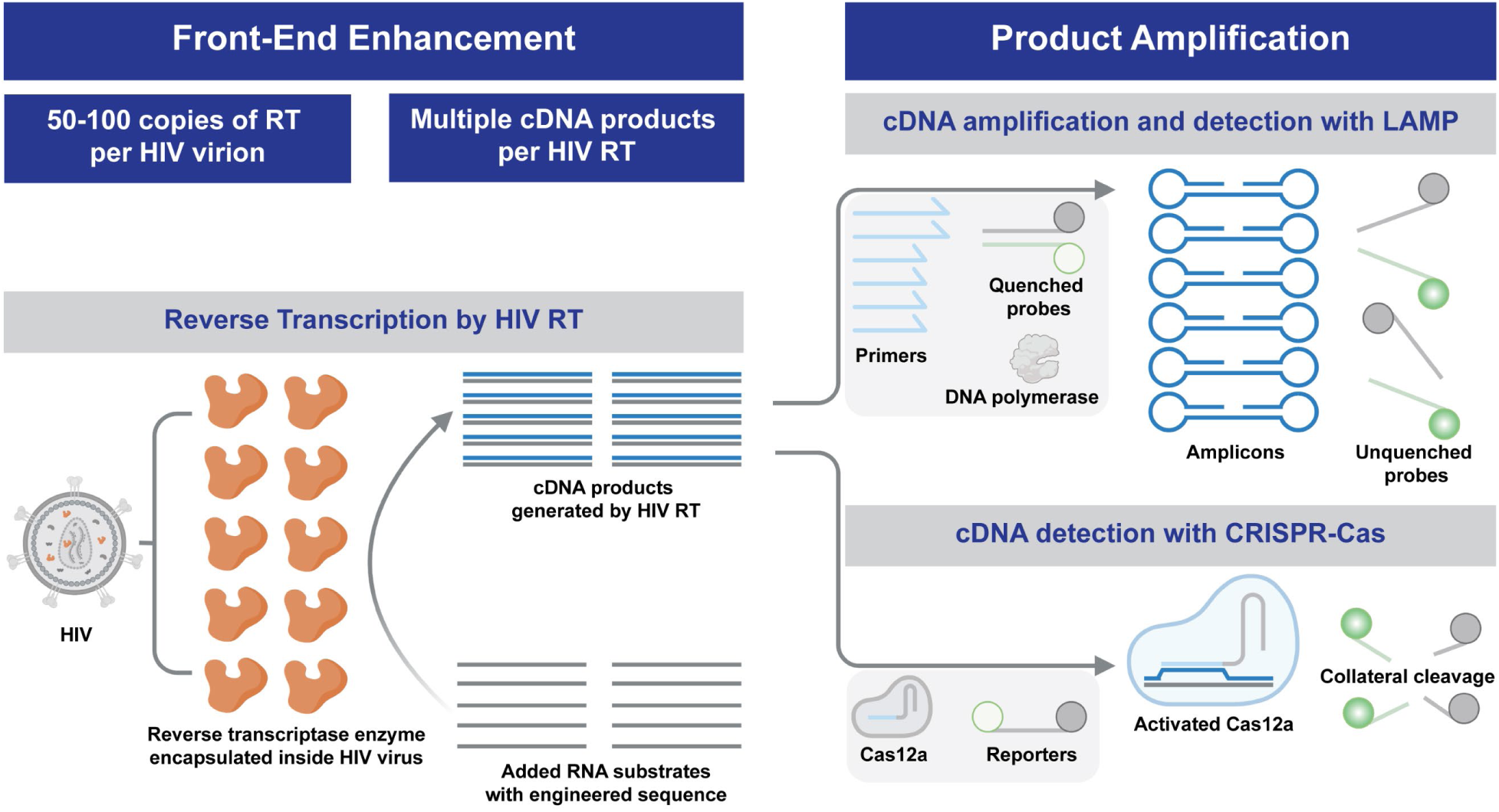
Illustrations of the proposed LAMP-mediated and Cas-mediated product-amplified reverse transcriptase activity assay.

Despite its superior sensitivity, LAMP does not provide direct quantification of the cDNA product by HIV RT enzyme and is thereby not applicable for phenotypic characterization. In recent years, the CRISPR-Cas system has emerged as a prominent tool for nucleic acid detection, where the CRISPR RNA (crRNA) recognizes the target sequence, subsequently activating the Cas enzyme and inducing collateral cleavage of reporters [36–39]. The CRISPR-Cas system offers linear signal amplification from activated Cas enzyme, suitable for quantification of the cDNA product and phenotypic characterization of HIV RT enzyme. Here we developed a Cas-mediated Product-Amplified Reverse Transcriptase activity assay (CasPART), where the crRNA-Cas12a was programmed to activate upon recognition of the target strand generated by HIV RT enzyme (Schematic 1). The high specificity of CRISPR-Cas towards the target strand enabled combining all assay reagents into one pot. We first optimized the substrate design to achieve higher sensitivity. Subsequently, we combined CasPART with direct immunocapture for sample preparation and demonstrated detection of HIV RT enzyme spiked into HIV-negative plasma. Furthermore, we explored using CasPART to report the response of wild-type and mutant HIV RT enzyme to antiretroviral drugs for potential drug resistance testing.

With its high programmability, CRISPR-Cas detection systems can be readily adapted to target other pathogen-derived enzymes by aligning the substrate architecture with the targeted enzyme activity. As a proof-of-concept, we further adapted CasPART to target the activity of HIV integrase (IN). With tailored substrate design, the crRNA-Cas12a system successfully detected the product of HIV IN in one pot, demonstrating its potential for broader applications. We anticipate that the approach of detecting enzyme products with CRISPR-Cas systems can be applied for phenotypic characterization of various pathogen-derived enzymes (Table 1), presenting a new avenue for drug development and phenotypic drug resistance testing.

## Results and Discussion

### HIV RT enzyme activity characterization with qPCR

A single copy of HIV RT enzyme can potentially generate multiple copies of cDNA, essentially providing a front-end enhancement. To guide the design of the RT activity assay and maximize the detection sensitivity, we first used qPCR to characterize the cDNA generation ability of HIV RT enzyme. We adapted previously reported protocols and made RNA substrates through transcription from DNA templates [40]. HIV RT enzyme was then incubated with the RNA substrates and other reagents at 37℃ for 2hr for cDNA generation. After heat inactivation, the sample was diluted and added to qPCR to quantify the copy number of cDNA generated by HIV RT enzyme (Figure 1A). The amplification from no-RT samples likely resulted from residual DNA templates in the added RNA substrates or endogenous reverse transcription activity of the OneTaq DNA polymerase used in qPCR [41]. All the positive samples with HIV RT enzyme showed significantly lower Cq compared to no-RT controls. As shown in Figure 1B, the copy number of cDNA and HIV RT enzyme exhibited a strong linear correlation, suggesting consistent cDNA per RT enzyme across a range of RT copy numbers. Moreover, one HIV RT enzyme generated 127.8 cDNA copies within 2hr, confirming the front-end enhancement from the catalytic activity of HIV RT enzyme. We next investigated the dependence of cDNA per HIV RT enzyme on incubation time and RNA substrate concentrations. The cDNA copy number per HIV RT enzyme decreased linearly when the incubation time was shortened from 2hr to 30min (Figure 1C and Supplementary Figure S1). A 30-minute incubation was thereby selected to balance the assay time and front-end enhancement. The cDNA copy number per HIV RT enzyme increased with increasing RNA substrate concentrations with saturation at 2nM and above (Figure 1D and Supplementary Figure S2). 5nM RNA was chosen to ensure the maximal front-end enhancement.

**Figure 1.**
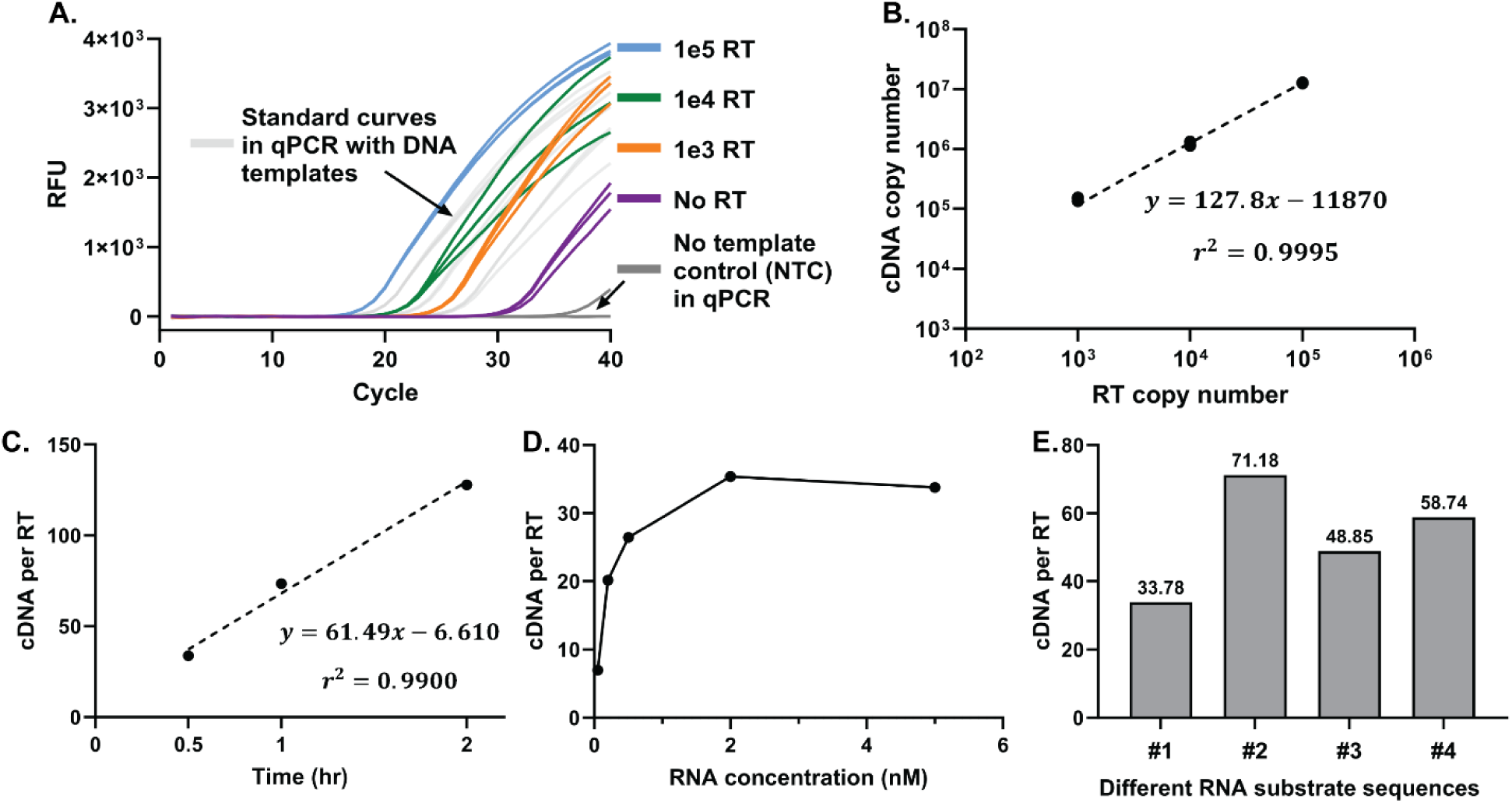
Characterization of HIV RT enzyme activity with qPCR. (A) Real-time curves of qPCR quantifying the copy number of cDNA generated by HIV RT enzyme with 2hr incubation and 5nM RNA substrates. (B) Linear correlation between the copy number of cDNA and copy number of HIV RT enzyme. The slope of the fitted line was used to represent the average cDNA copy number per HIV RT enzyme. (C) The cDNA per HIV RT enzyme with different incubation time and 5nM RNA substrates. Linear correlation of cDNA per RT enzyme at each data point can be found in Supplementary Figure S1. (D) The cDNA per RT enzyme with different concentrations of RNA substrates and 0.5hr incubation. Linear correlation of cDNA per RT enzyme at each data point can be found in Supplementary Figure S2. (E) The cDNA per RT enzyme on different RNA substrate sequences with 0.5hr incubation and 5nM RNA. Sequence #1 was used in panel A-D. Linear correlation of cDNA per RT enzyme for sequences #2, #3, and #4 with 0.5nM, 2nM, and 5nM RNA substrates can be found in Supplementary Figure S3. All linear correlations were conducted in GraphPad Prism 10.

When transitioning to LAMP for cDNA amplification, we selected multiple RNA substrates of engineered sequences with established LAMP assays developed within our group (Supplementary Information Sequence Table). Interestingly, we found that the cDNA copy number per HIV RT enzyme varied among the different RNA sequences (Figure 1E and Supplementary Figure S3). The differences may be caused by secondary structures of the RNA substrates that led to varied reverse transcription efficiency, or sequence preference of HIV RT enzyme as reported previously [42]. Nevertheless, sequence #2 with the highest cDNA copy number per HIV RT enzyme was selected for assay development aiming for high sensitivity.

### Development of one-pot LamPART assay

We aimed to combine cDNA generation by HIV RT enzyme and subsequent cDNA amplification by LAMP into one pot to simplify the assay workflow. Specifically, the reaction included the reagents for both reverse transcription and LAMP and simply incubated at 37℃ for 30 minutes for cDNA generation followed by 65℃ for 1 hour for LAMP. The LAMP assay employed previously developed sequence-specific probes to report amplification [43]. However, we initially encountered strong false positives in the absence of HIV RT enzyme, likely caused by innate reverse transcription activity of the DNA polymerase in LAMP at both 37℃ and 65℃ [44], and/or side products from primer dimers during 37℃ incubation that led to nonspecific amplification in LAMP (Figure 2A and 2C). TFpol, a chimeric polymerase variant previously developed within our group, was used as the LAMP DNA polymerase since it was reported to be highly specific [35,43]. In addition, TFpol showed less reverse transcription activity compared to Bst 2.0 WS as quantified by qPCR (Supplementary Figure S4). However, TFpol still generated over 10^5^ and 10^10^ copies of cDNA when incubated with 5nM RNA for 30min at 37℃ and 65℃, respectively (Supplementary Figure S4). To address the issue, first, an aptamer against the backbone of TFpol was included in the reaction to inhibit TFpol activity at 37℃ [45,46]. Second, to differentiate the cDNA product from the RNA substrate, the RNA sequence was truncated to be shorter than the LAMP footprint and missing F1 and partial F2 primer binding regions (Figure 2B). At 37℃, the FIP (F1-F2) primer functioned as the RT primer and hybridized with the truncated RNA substrate to generate the full-length cDNA driven by HIV RT enzyme. The melting temperature of the hybridization region was designed at around 45℃. At 65℃, any unextended FIP-RNA hybrid dissociated, and the truncated RNA substrate was non-amplifiable in LAMP, eliminating false positives from innate reverse transcription activity of TFpol. Only the full-length cDNA generated by HIV RT enzyme could be amplified and detected. In addition, the cDNA had one loop structure incorporated, removing the need for the F3 primer and one bumping step in LAMP that was suggested to be rate-limiting [47]. Furthermore, to address the issue of nonspecific amplification from primer dimers generating side products at 37℃, we introduced dockers to LAMP primers, specifically to the LB and B3 primers. The dockers are short DNA oligos complementary to the matching primer with melting temperatures around 55℃. The dockers could sequester the LB and B3 primers at 37°C to prevent extendable primer dimers and then release the primers at 65℃ for LAMP amplification (Figure 2B). The dockers were blocked on the 3’ end to avoid introducing new extendable dimers. Besides assay designs to suppress false positives, interestingly, we also found that cDNA per HIV RT enzyme under LAMP conditions started to saturate at 50nM RNA instead of 2nM (Supplementary Figure S5). The discrepancy could be attributed to much higher primer concentrations in LAMP and primer dimers at 37℃ competing against RNA substrates to bind HIV RT enzyme, or higher dNTP concentrations in LAMP that chelated Mg^2+^ and thereby affected HIV RT enzyme activity. Regardless, 50nM RNA substrate was used in LamPART for a balance between HIV RT efficiency and false positives from TFpol acting on RNA substrates.

**Figure 2.**
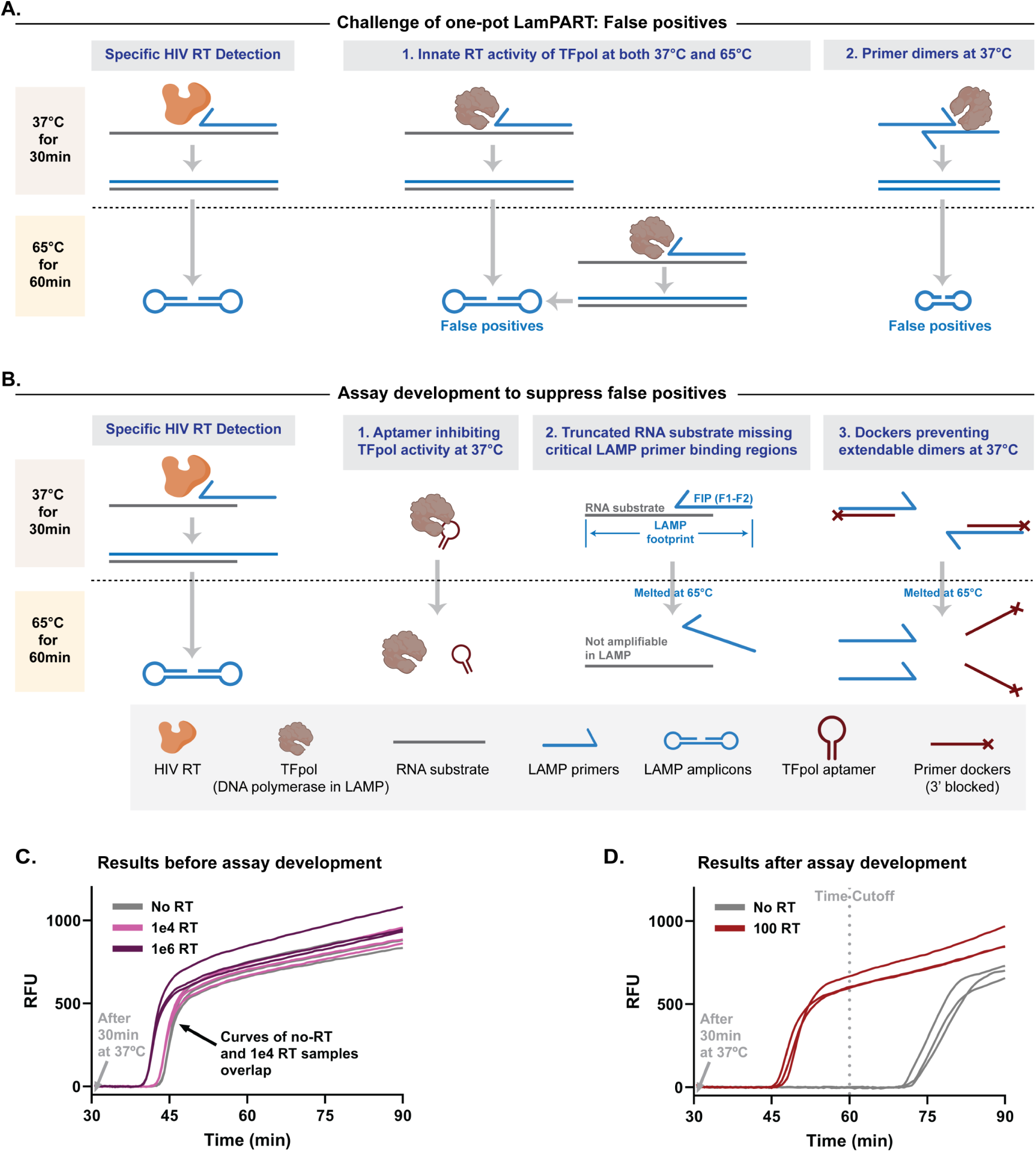
Development of one-pot LamPART assay. (A) Illustrations of the false positive challenge in one-pot LamPART assay. (B) Assay development employed to suppress false positives. (C) One-pot LamPART assay results before assay development. (D) One-pot LamPART assay results after assay development.

Incorporating all these developments in the one-pot LamPART assay, we successfully suppressed the amplification of the negative controls to lift off after 60 minutes (30 minutes at 37℃ and 30 minutes at 65℃), while 100-RT samples lifted off before 60 minutes (Figure 2D). The 60-minute duration was thereby established as the assay cutoff time. Compared to previous HIV RT enzyme activity assays that require separate steps involving cDNA generation, RNA substrate digestion, and cDNA amplification, the one-pot LamPART assay greatly simplified the workflow due to the novel assay designs. The whole assay is within an hour and requires only two temperature steps without user interventions in between. Moreover, we introduced the concept of primer dockers that could sequester unwanted primers at lower temperatures to prevent primer dimers and side products, while releasing the primers at higher temperatures for their proper function. The design of primer dockers can be applied to other molecular assays involving multiple temperatures, to minimize unspecific reactions and improve assay performance.

### Performance of the one-pot LamPART assay

We then characterized the limit of detection (LoD) of the one-pot LamPART assay on HIV RT enzyme. In preliminary LoD screening, the assay successfully detected all three replicates of 10-RT samples. In further LoD determination with 20 replicates, while only 11 replicates were positive for 10-RT samples, 19 replicates of 20-RT samples showed positive (Figure 3A and Supplementary Figure S6). All 20 replicates of negative controls were amplified later than the cutoff time. Therefore, the one-pot LamPART assay demonstrated a LoD at 20 copies of recombinant HIV RT enzyme, equivalent to a single virion.

**Figure 3.**
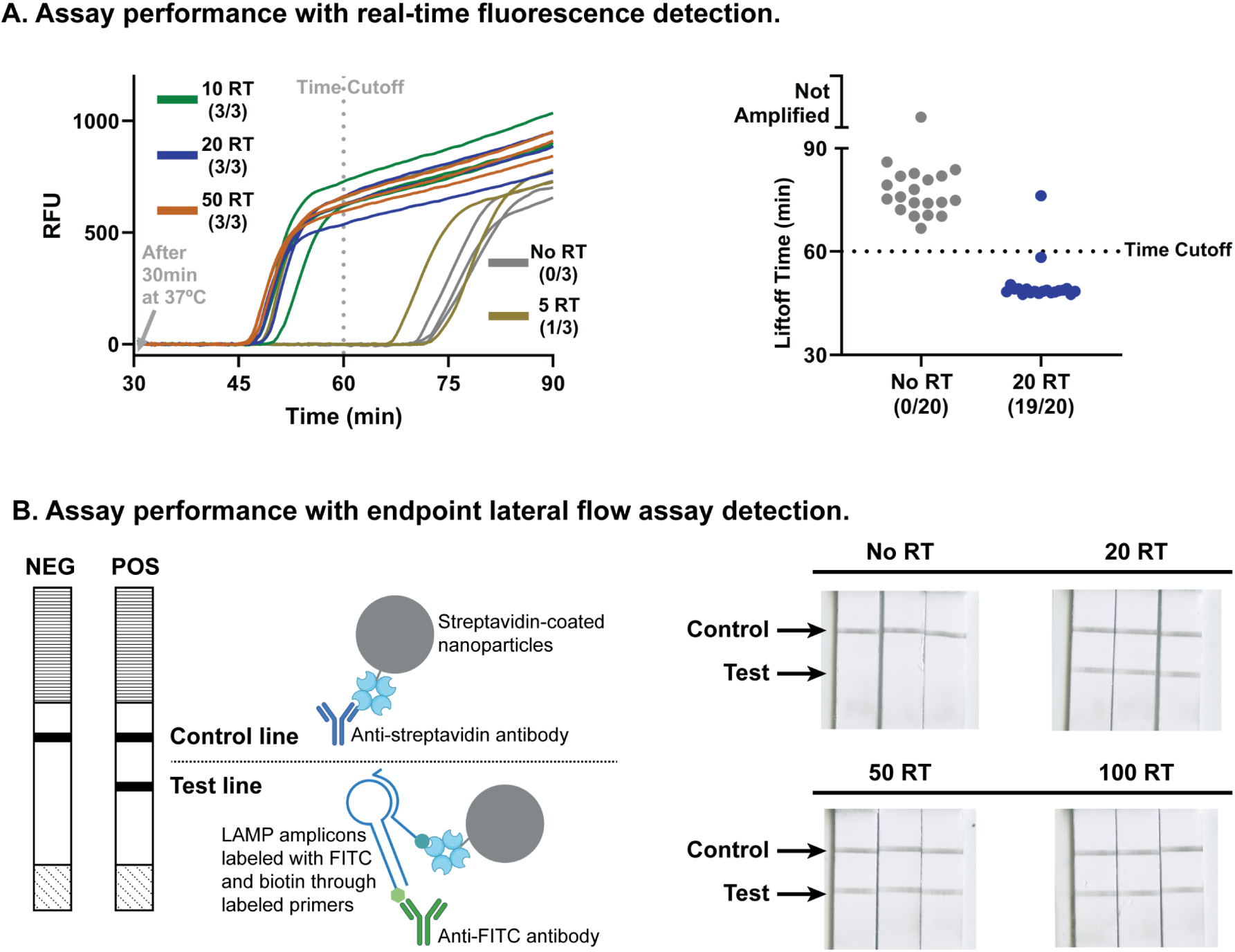
One-pot LamPART assay performance. (A) Performance of the one-pot LamPART assay with real-time fluorescence detection. Left, preliminary LoD screening with three replicates per sample. Right, LoD determination with 20 replicates of negative and 20-RT samples. The results for 20 replicates with 10-RT samples can be found in Supplementary Figure S6. (B) Performance of the one-pot LamPART assay with endpoint lateral flow assay detection. Left, schematics illustrating the compositions at the control line and test line. Right, scanned images of the lateral flow strip detection region. A detailed illustration of the lateral flow assay detection workflow can be found in Supplementary Figure S7.

To further reduce the instrument complexity, we developed endpoint lateral flow assay (LFA) for LAMP detection to remove the need of real-time fluorescence measurement. We adapted a published method where LAMP primers FIP and BIP were labeled with biotin and FITC, respectively [48]. The LAMP amplicons were dual-labeled and subsequently detected using a commercial lateral flow strip (Figure 3B and Supplementary Figure S7). The results showed that the test lines remained clean for negative controls and appeared for 20-RT, 50-RT, and 100-RT samples (Figure 3B). The one-pot LamPART assay with endpoint LFA detection demonstrated the same LoD as real-time fluorescence readout, 20 copies of recombinant HIV RT enzyme.

### LamPART-based integrated workflow from sample to detection

To build the integrated workflow, we developed direct immunocapture of HIV RT enzyme from plasma samples for sample preparation (Figure 4A). Specifically, fingerstick plasma was mixed with lysis buffer which included 0.6% NP-40 for virus lysis, adapted from previous HIV RT enzyme assays, and anti-HIV-RT-coated magnetic beads for HIV RT enzyme capture. The first step for development was to identify anti-HIV-RT antibodies that do not inhibit HIV RT enzyme activity. By using qPCR to compare cDNA generation between HIV RT enzyme preincubated with antibodies and free HIV RT enzyme, we selected an antibody that preserved the function of the HIV RT enzyme (Supplementary Figure S8).The antibody was then bound to anti-mouse-IgG-conjugated magnetic beads for immunocapture of HIV RT enzyme. To avoid false positives from unspecific absorption of human polymerases that may have reverse transcription activity [49], 1% BSA was used to pre-block the beads and added to the lysis buffer during HIV RT capture for additional real-time blocking. To evaluate the performance of the developed direct immunocapture method, we used qPCR to quantify cDNA generation from HIV RT enzyme spiked into HIV-negative plasma and subsequently captured. As shown in Figure 4B, the negative controls generated negligible cDNA, confirming the specificity of the immunocapture approach. The captured HIV RT enzyme generated around half the quantity of cDNA compared to the positive controls (HIV RT enzyme directly added into the assay). The loss could be due to imperfect capture efficiency or the inhibitory effect of beads on the activity of bound HIV RT enzyme. Regardless, the results validated the feasibility of the direct immunocapture approach.

**Figure 4.**
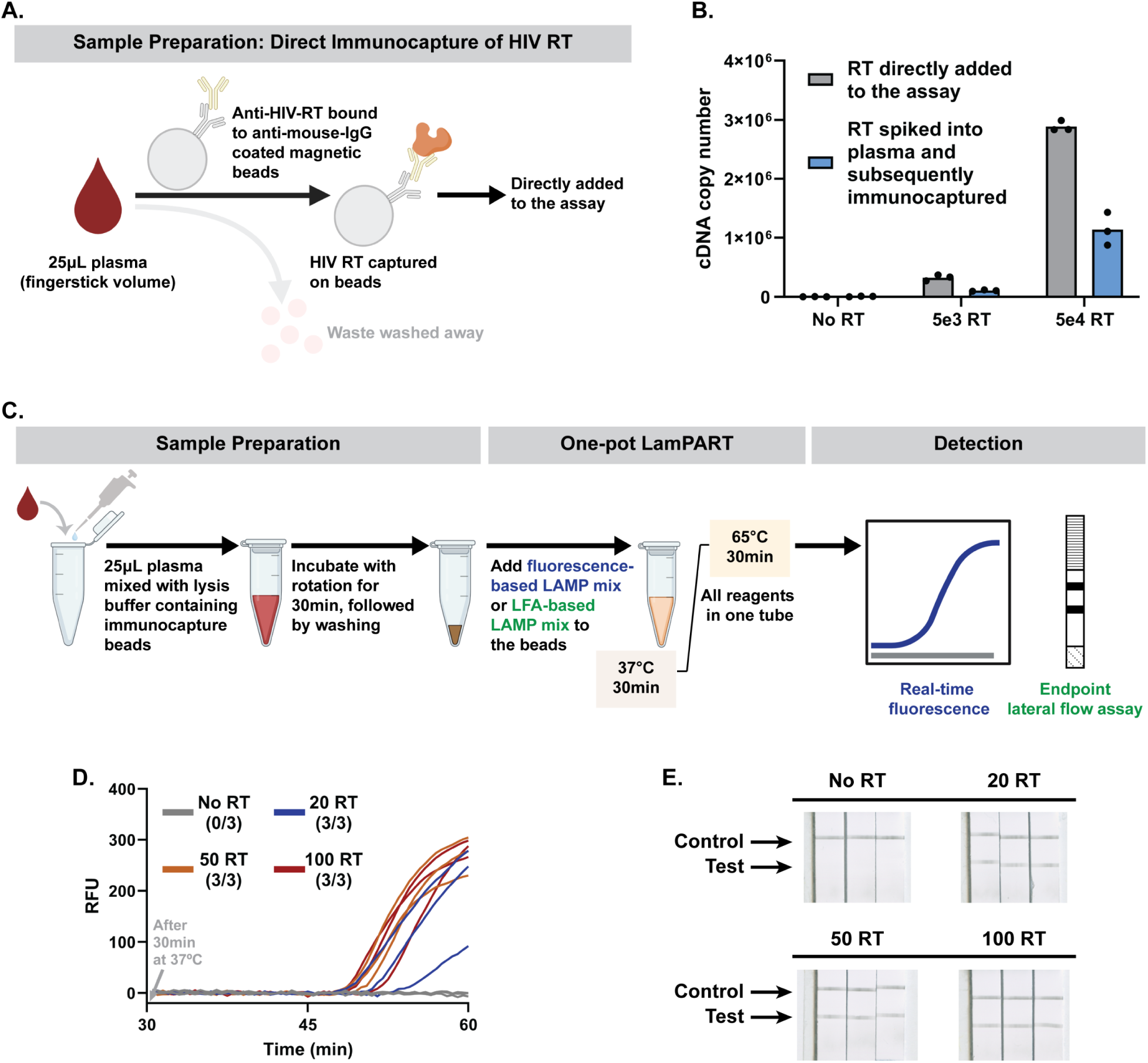
LamPART-based integrated workflow. (A) Schematics illustrating the sample preparation approach, direct immunocapture of HIV RT enzyme from plasma. (B) qPCR quantification of cDNA generation from HIV RT enzyme directly added to the assay versus HIV RT enzyme spiked into HIV-negative plasma and subsequently immunocaptured. (C) Illustration of the integrated workflow, with either real-time fluorescence detection or endpoint lateral flow assay detection. (D) and (E) Results of the integrated workflow with real-time fluorescence detection (D) or endpoint lateral flow assay detection (E) on 25μL HIV-negative plasma spiked with HIV RT enzyme.

Combining the sample preparation and the one-pot LamPART assay altogether, we developed the integrated sample-to-result workflow, where the bead-captured HIV RT enzyme was directly added to the assay with either real-time fluorescence or endpoint LFA detection (Figure 4C). We tested the workflow using HIV-negative plasma spiked with recombinant HIV RT enzyme. Both the fluorescence-based and LFA-based workflow successfully detected 20 copies of recombinant HIV RT enzyme spiked into 25μL HIV-negative plasma (Figure 4D and 4E), equivalent to a single virion. In addition, all the no-RT samples showed negative results, demonstrating the specificity of the workflow.

The integrated workflow demonstrated a LoD at 20 copies of recombinant HIV RT enzyme spiked into 25μL plasma (fingerstick volume), highlighting its potential for sensitive HIV detection at the POC settings. The whole workflow required less than a 2-hour total turnaround time, with minimal user steps necessitating less than 30 minutes of hands-on time. The instrumentation consisted solely of a magnetic stand and a thermocycler that could run two temperatures combined with endpoint lateral flow detection. In addition, the workflow could be conducted in a high throughput manner by employing PCR strip tubes for sample processing combined with multi-channel pipettes (Supplementary Figure S9). Several aspects of the workflow could be further developed to facilitate simpler implementation. First, the antibody capture and washing steps can be automated in microfluidic chips to further reduce user interventions and shorten the total turnaround time. Second, the RNA substrates currently need to be stored in -80℃ freezers, challenging for POC testing especially in low-resource settings. Future work could explore expressing the RNA substrates in phage-like particles to allow storage at 4℃ or even ambient temperatures [50]. Lastly, the assay reagents could be lyophilized and rehydrated at the point of testing to further simplify the reagent storage and assay setup steps [51]. Nevertheless, the high sensitivity and simple procedures of the LamPART-based integrated workflow provided an alternative and promising option for sensitive HIV testing at the POC.

### Development of one-pot CasPART assay

CRISPR-Cas systems have been widely used in recent years for nucleic acid detection, with high specificity towards the target strand and linear signal amplification from the nuclease activity of activated Cas enzymes. Here, we aimed to develop CasPART assay where the target strand generated by HIV RT enzyme could specifically activate the crRNA-Cas12a complex, to unleash DNase I activity and cut reporters for fluorescence generation (Figure 5A). The proposed CasPART assay presents several advantages. First, the high specificity of crRNA-Cas12a facilitates the inclusion of all assay reagents, including the substrates for HIV RT enzyme, into a one-pot reaction. In addition, the optimal temperature for crRNA-Cas12a is 37°C, same as the reaction temperature for HIV RT enzyme, allowing a constant temperature for the one-pot CasPART assay. Moreover, the linear signal amplification of crRNA-Cas12a detection allows quantifying the products generated by HIV RT enzyme and thereby characterizing the HIV RT enzyme activity. Since HIV RT enzyme is a common target for antiretroviral therapy (ART), the phenotypic characterization capability of CasPART assay holds promise for HIV drug resistance testing and therapeutic drug monitoring.

**Figure 5.**
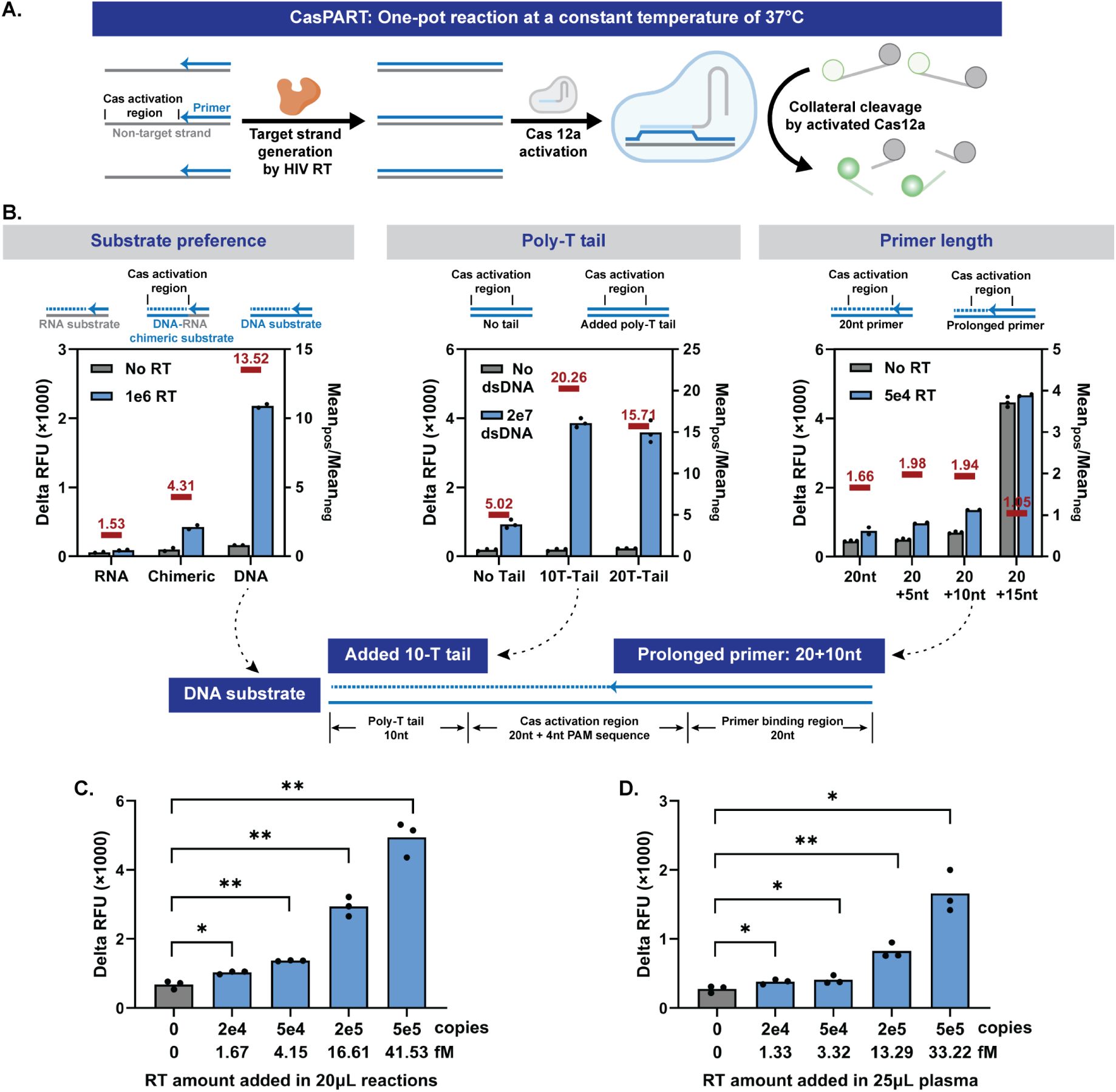
Development of the CasPART assay. (A) Illustrations of the one-pot CasPART assay. (B) Designs of the HIV RT substrate in CasPART to improve assay sensitivity. Delta RFU during 2hr incubation (endpoint RFU minus starting RFU) was used as assay signals. Left, CasPART signals with DNA, DNA-RNA chimeric, and RNA substrates. Middle, assay signals of incubating crRNA-Cas12a with double-stranded DNA with or without poly-T tails next to the activation region. Right, CasPART signals with varied primer lengths in the substrate. Bottom, illustrations of the final substrate design. (C) Performance of CasPART with HIV RT enzyme directly added in the assay. Real-time curves of 5hr incubation and delta RFU during 0.5hr and 5hr incubation can be found in Supplementary Figure S11. (D) Performance of CasPART integrated with direct immunocapture for sample preparation on 25μL HIV-negative plasma spiked with HIV RT enzyme. Statistical significance in panels C and D was determined by unpaired t tests with Welch’s correction and conducted in GraphPad Prism 10. * indicates p < 0.05 and ** indicates p < 0.01.

Due to limited signal amplification from activated Cas enzymes, previously reported CRISPR-Cas detection systems often require pre-amplification of the target nucleic acid to achieve clinically relevant sensitivity. In CasPART, however, the front-end enhancement offered by HIV RT enzyme could inherently improve the assay sensitivity. Regardless, we started to develop the one-pot CasPART assay aiming for low LoD. First, since Cas12a prefers double-stranded DNA substrates over DNA-RNA hybrid and HIV RT enzyme also has DNA-dependent DNA polymerization activity [25,52], we investigated whether RNA or DNA substrates in CasPART could achieve lower LoD. We started with the RNA substrate sequence #1 in Figure 1E and picked a 20nt region with adjacent PAM sequence and minimal hairpin structures as the crRNA-Cas12a target. The same sequence plus PAM and primer binding region was used to create the DNA substrate. The fluorescence increase (endpoint RFU minus starting RFU) during 2-hour incubation was used as the assay readout. As shown in Figure 5B, the DNA substrates generated much higher positive signals. Interestingly, the background signals were lower with RNA substrates, likely due to less unspecific activation of crRNA-Cas12a. Therefore, we further tested DNA-RNA chimeric substrates, where the Cas12a activation region was DNA and the remaining sequence was RNA, aiming for high positive signals while maintaining low background signals. With chimeric substrates, while the background signals stayed low, the positive signals were still much lower than the DNA substrates. Therefore, we selected DNA substrates to move forward. Although DNA substrates could be extended by the endogenous human DNA polymerase in patient samples and pose risks of false positives, the developed method of immunocapturing HIV RT enzyme for sample preparation (Figure 4A) ensures the specificity of the assay. Besides, DNA substrates are more stable for storage and simpler for implementation. Next, we investigated if the crRNA-Cas12a activation requires a substrate footprint longer than the activation region alone. We tested incubating crRNA-Cas12a with double-stranded DNA, representing the products generated by HIV RT enzyme, with or without a poly-T tail next to the activation region. Interestingly, substrates with 10-T or 20-T tails exhibited significantly higher signals compared to those with no tails (Figure 5B), indicating support for the crRNA-Cas12a activation from regions adjacent to the activation sequence. A 10-T tail was therefore added in the substrate design. Additionally, we tested including two Cas12a activation regions in the substrate to potentially activate more Cas12a per HIV RT enzyme product for signal enhancement, a strategy reported in Cas13a nucleic acid detection systems [53]. However, substrates with dual activation regions produced weaker signals and were thereby not adopted in the substrate design (Supplementary Figure S10). Lastly, we tested increasing the primer length to minimize the HIV RT extension required for Cas12a activation to improve assay sensitivity. The positive signals increased as primers spanned into the activation region, but the background signals rose sharply when the overlap of the primer and the activation region changed from 10nt to 15nt, indicating a tipping point in the target strand for nonspecific Cas12a activation (Figure 5B). The 20+10nt primer was thereby employed in the final assay.

Incorporating all the developments, we characterized the LoD of the one-pot CasPART assay. As shown in Figure 5C, the assay with 2hr incubation detected down to 2e4 copies (1.67fM) of HIV RT enzyme directly added to the reaction. When combined with the direct immunocapture method for sample preparation, CasPART demonstrated the same LoD at 2e4 copies of HIV RT enzyme spiked into 25μL plasma (1.33fM) (Figure 5D), while the negative controls generated low background signals thanks to the highly-specific sample preparation method. The developed CasPART assay, a one-pot reaction at a constant temperature of 37°C, demonstrated femtomolar sensitivity for protein detection, which is among the highest in immunoassays [54].

### CasPART assay for phenotypic characterization of HIV RT enzyme

We next explored the capability of CasPART to characterize HIV RT enzyme activity under antiretroviral drugs used for HIV treatment. The CasPART assay signal should decrease in the presence of antiretroviral drugs (Figure 6A), holding potential for measuring drug concentrations in patient samples for therapeutic drug monitoring. Moreover, with the phenotypic characterization capability, the CasPART assay can distinguish the different responses of wild-type (WT) and mutant (MUT) HIV RT enzymes to antiretroviral drugs, making it promising as a phenotypic drug resistance test.

**Figure 6.**
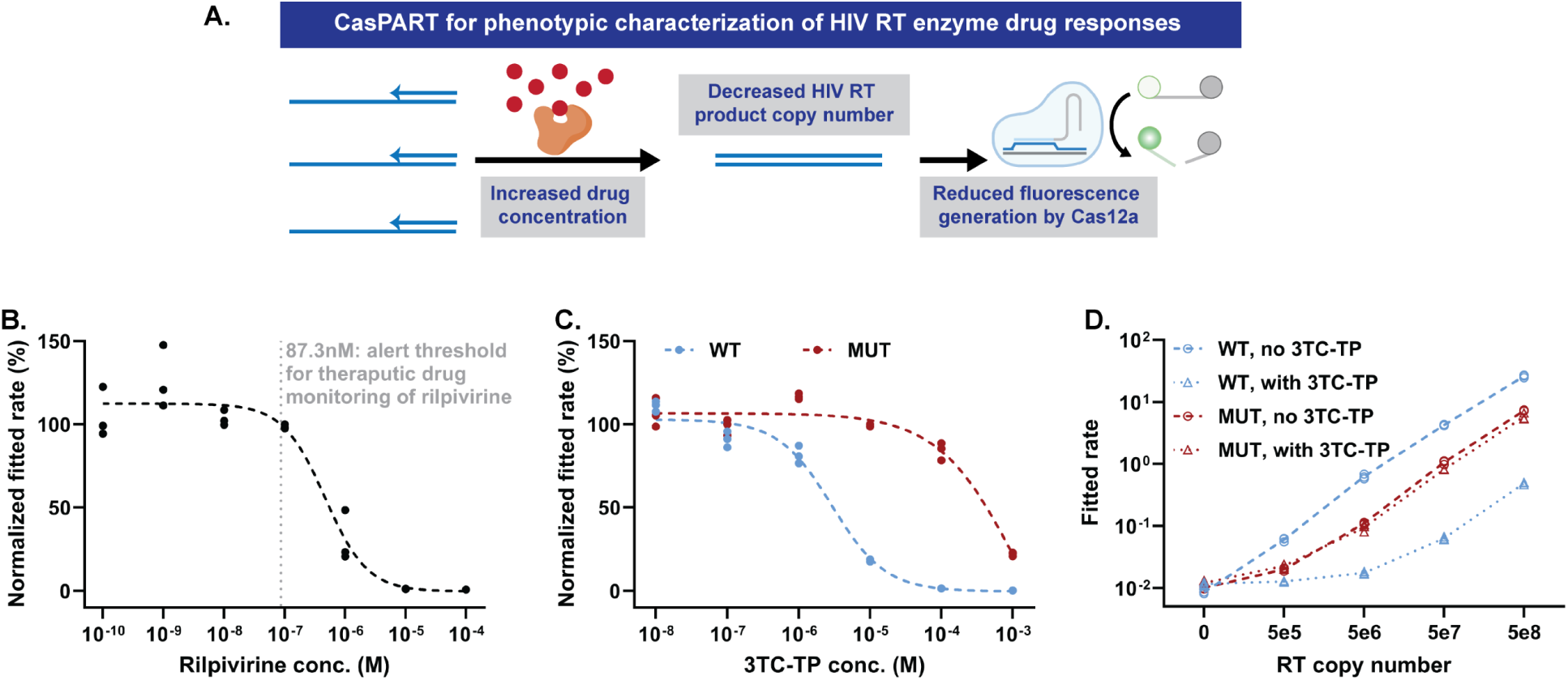
CasPART for phenotypic characterization of HIV RT enzyme. (A) Illustrations of the phenotypic characterization capability of CasPART. (B) The CasPART response of HIV RT enzyme to the antiretroviral drug rilpivirine for potential therapeutic drug monitoring. The HIV RT enzyme copy number was set at 1e7 in all samples. Assay signals from samples with rilpivirine were normalized against the average of the three control samples without rilpivirine. The dashed curve was fitted four-parameter logistic regression curve using GraphPad Prism 10. The dotted vertical line refers to the alert threshold of rilpivirine concentration in patient samples recommended for therapeutic drug monitoring [57]. The real-time curves can be found in Supplementary Figure S12. The fitted rates of all samples can be found in Supplementary Table S1. (C) The CasPART response of wild-type (WT) and mutant (MUT) HIV RT enzyme to the antiretroviral drug 3TC-TP. The HIV RT enzyme copy number was set at 1e7 in all samples. Assay signals from samples with 3TC-TP were normalized against control samples without 3TC-TP. The dashed curve was fitted four-parameter logistic regression curve using GraphPad Prism 10. The real-time curves can be found in Supplementary Figure S14. The fitted rates of all samples can be found in Supplementary Table S2 and S3. (D) The fitted rates of WT and MUT HIV RT enzyme at different copy numbers with or without 10^-4^ M 3TC-TP. The real-time curves can be found in Supplementary Figure S15. The fitted rates of all samples can be found in Supplementary Table S4, S5, S6, and S7.

We first tested using CasPART for therapeutic drug monitoring. Measuring antiretroviral drug concentrations in patient samples can enable identification of interindividual differences in drug levels based on missed doses, drug-drug interactions, or differences in drug pharmacokinetics due to age, pregnancy, or body size [55,56]. As a proof-of-concept, we tested measuring rilpivirine, a non-nucleoside reverse transcriptase inhibitor (NNRTI) used in ART regimens. To avoid endpoint fluorescence plateaus in CasPART, we fitted the real-time fluorescence curve to a second order polynomial equation and used the quadratic coefficient as the fitted rate for HIV RT enzyme (the fitting details are described in the method section). The fitted rates from samples with different concentrations of rilpivirine (10^-10^ – 10^-4^M) were normalized against control samples with no rilpivirine. As expected, assay signals were not inhibited with low rilpivirine concentrations but decreased rapidly with higher concentrations, with a 50% inhibition concentration (IC_50_) at 462nM (Figure 6B and Supplementary Figure S12). The results demonstrated that CasPART held potentials in distinguishing rilpivirine concentrations above or below the alert threshold (87.3nM) for therapeutic monitoring of rilpivirine recommended by the French National AIDS Agency to avoid treatment failure [57]. In addition to therapeutic drug monitoring, CasPART with the demonstrated ability to characterize drug inhibition could potentially be employed for the development of antiretroviral drugs.

We further investigated the CasPART responses of WT and MUT HIV RT enzyme to lamivudine triphosphate (3TC-TP), a nucleoside RT inhibitor (NRTI) often included in ART regimens, for potential applications in phenotypic HIV drug resistance testing. The MUT RT carried an M184V mutation known to present resistance to 3TC-TP [58]. Since 3TC-TP is an analog to dCTP, we first tested decreasing dCTP concentrations in CasPART, to favor the competition with 3TC-TP, a strategy employed in previous reports [59]. 10μM dCTP was selected as it was the lowest concentration that did not compromise CasPART signals (Supplementary Figure S13). Next, we tested CasPART signal inhibition using 1e7 copies of WT or MUT RT across varying concentrations of 3TC-TP (10^-8^ – 10^-3^M), to observe their responses to 3TC-TP and determine the optimal 3TC-TP concentration for drug resistance testing. The WT and MUT RT exhibited substantially different 3TC-TP responses, with MUT RT requiring approximately a 100-fold higher 3TC-TP concentration for inhibition compared to WT RT. The results highlighted the capability of CasPART to differentiate between WT and MUT RT (Figure 6C and Supplementary Figure S14). A concentration of 100μM 3TC-TP, resulting in nearly complete inhibition of WT RT with minimal impact on MUT RT, was therefore selected for drug resistance testing. Finally, with the identified conditions, we tested CasPART signals across a range of WT or MUT RT copy numbers with or without 3TC-TP, as a proof-of-concept for phenotypic drug resistance testing. As shown in Figure 6C and Supplementary Figure S15, signals from WT RT across 5e5 to 5e8 copies were greatly reduced in the presence of 3TC-TP, while MUT RT signals remained similar, demonstrating successful detection of MUT RT. Interestingly, the MUT RT generated lower signals in CasPART compared to WT, which could be due to lower purity of the MUT RT stock from a different source or reduced overall RT activity induced by the drug-resistance mutation [60]. Nonetheless, the results demonstrated the ability of CasPART to detect drug resistance in HIV RT enzyme, providing a phenotypic measure of drug resistance that has potential to be applied for minimally instrumented POC screening.

Although this initial demonstration of drug resistance testing is promising, multiple issues need to be addressed in future work to enable clinical implementations. First, the sensitivity of the current CasPART assay do not cover lower HIV viral loads among patients using fingerstick samples. The issue could potentially be resolved by using larger sample volumes (≥100 µL) that can be collected with user-friendly devices such as the Tasso blood collector [61]. Second, the efficiency of immunocapture with MUT HIV RT enzyme needs to be tested. Last, the overall lower activity of MUT HIV RT compared to WT may pose challenges in detecting mutants mixed with wild type. However, prior HIV RT activity assays with PCR-based readout showed that as low as 10% mutant mixed with wild type HIV RT can be detected [62]. The substrate sequence in CasPART could also be systematically designed to match the targeted NRTI drug, to maximize enzyme inhibition events and potentially further favor the detection of MUT RT [56]. Future work is needed to investigate the sensitivity of CasPART in detecting the MUT proportion and optimize assay conditions such as drug concentrations accordingly. Regardless, the CasPART assay demonstrated potential for phenotypic drug resistance testing, especially in low-resource settings where other tests are not available.

### Adapting CasPART for other enzyme targets

Besides RT enzyme, HIV also carries integrase with unique activity. Beyond HIV, other pathogens also possess various enzymes with specialized activities (Table 1). The crRNA-Cas12a detection system, with its highly programmable target sequence and superior specificity towards the target strand, can be readily adapted to detect other pathogen-derived enzymes by tuning the substrate architecture to match the targeted enzyme activity. As a proof-of-concept, we adapted CasPART to detect HIV integrase (IN). HIV IN exhibits disintegration activity in vitro, where the 3’ hydroxyl group at the disintegration site attacks the adjacent phosphodiester bond, generating the linked product (Figure 7A) [63,64]. Unlike the integration activity of HIV IN which produces random products in vitro [65,66], disintegration generates specific products and is thereby selected as the assay target. We adapted the previously reported substrate design and changed the sequence to match the crRNA-Cas12a used in CasPART [63]. Since the substrate contained the whole sequence targeted by crRNA-Cas12a and only missed a single phosphodiester bond at the disintegration site, the biggest challenge was the background signal caused by nonspecific activation of Cas12a. To address the issue, we varied the distance between the disintegration site and the PAM region on the substrate and found that a 10nt distance reduced the background signals to a level similar to the CasPART (Figure 7B). Interestingly, substrates with 0nt, 5nt, and 20nt distances generated similarly high background signals, suggesting that the 10nt position may be a tipping point for nonspecific Cas12a activation, aligning with the results of varied primer lengths in CasPART (Figure 5B). As shown in Figure 7C, the final assay with the 10nt-distance substrates showed a LoD at 1e12 copies (83nM) of HIV IN. The slightly decreased signals at 1e13 IN copies were likely caused by inhibition from the storage buffer of the IN stock. The overall high LoD was possibly due to the slow activity of HIV IN, as quantified by qPCR (Supplementary Figure S16) and reported in the literature [65]. Nevertheless, the results demonstrated successful adaptation of CasPART for phenotypic detection of other enzymes for broader applications. The specificity of the assays can be further secured by the immunocapture of the targeted enzyme or the whole pathogen during sample preparation.

**Figure 7.**
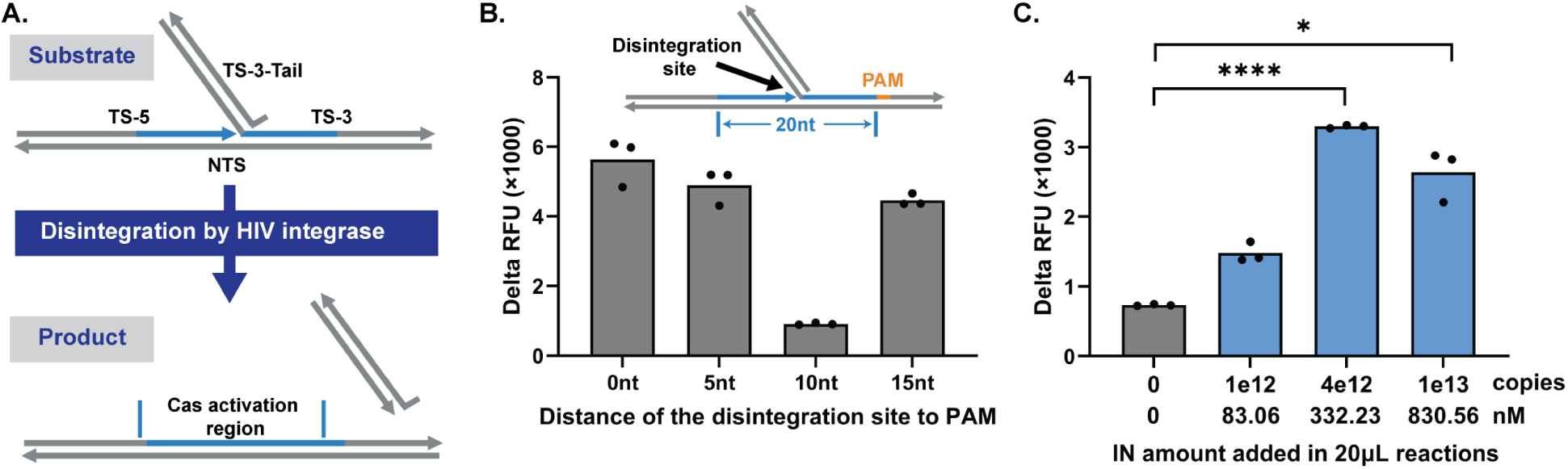
Adapting CasPART for other enzyme targets. (A) Illustrations of the disintegration activity of HIV IN. Under HIV IN catalysis, the 3’ hydroxyl group of TS-5 attacks the adjacent phosphodiester bond in TS-3, generating the linked product as the target strand for crRNA-Cas12a detection. (B) Background signals of using substrates with different distances between PAM and the disintegration site. (C) The final performance of detecting HIV IN using crRNA-Cas12a. Delta RFU during 2hr incubation was plotted in panel B and C. Statistical significance in panel C was determined by unpaired t-tests with Welch’s correction and conducted in GraphPad Prism 10. * indicates p < 0.05 and **** indicates p < 0.0001.

## Conclusions

We aimed to develop an HIV RT enzyme activity assay for sensitive HIV detection and phenotypic characterization due to several advantages of RT enzyme as a biomarker, including higher copy number per virion, multiple cDNA products per RT enzyme, robustness in plasma samples, conserved activity across subtypes, and offering direct measure of the antiretroviral drug efficacy. The cDNA generation activity of HIV RT enzyme was first characterized with qPCR to guide assay designs. We found that cDNA per RT enzyme appeared linear over time, saturated at ∼2nM RNA substrates, and varied among different substrate sequences. The sequence with the highest RT efficiency was selected for assay development, where one RT enzyme generated 71.18 copies of cDNA within 30 minutes, providing a front-end enhancement.

Based on the characterization of HIV RT enzyme activity, two formats of HIV RT enzyme activity assay, LamPART and CasPART, were then developed. In LamPART, LAMP was used to amplify cDNA products generated by HIV RT enzyme. With novel assay designs, we achieved a one-pot reaction with 30min incubation at 37°C followed by another 30min at 65°C. The assay demonstrated a LoD of 20 copies of recombinant HIV RT enzyme, equivalent to a single virion. The one-pot LamPART assay was further integrated with direct immunocapture of HIV RT enzyme using antibody-coated magnetic beads and endpoint LFA detection of LAMP amplicons. The integrated workflow achieved detection of 20 copies of recombinant HIV RT enzyme spiked into 25μL HIV-negative plasma samples.

In CasPART, we combined the front-end enhancement by HIV RT enzyme and the specific target strand detection by the CRISPR-Cas system, enabling one-pot reaction at a constant temperature of 37°C. We tailored the substrate design to improve assay sensitivity. The final assay, when combined with direct immunocapture for sample preparation, demonstrated a LoD of 2e4 copies (1.33fM) of recombinant HIV RT enzyme spiked into 25μL plasma. Furthermore, the linear signal amplification from CRISPR-Cas enabled CasPART as a phenotypic assay to characterize HIV RT enzyme activities. We used CasPART to measure the inhibition of HIV RT enzyme activity from rilpivirine, an antiretroviral drug, with potential applications in therapeutic drug monitoring. In addition, CasPART successfully differentiated wild-type HIV RT enzyme from mutants with known resistance to 3TC-TP, another drug commonly included in ART regimens, demonstrating the potential of CasPART for phenotypic drug resistance testing.

Moreover, the inherent programmability of the CRISPR-Cas system allowed simple adaptation of CasPART for other pathogen-derived enzymes. As a proof-of-concept, we adapted CasPART to detect the disintegration activity of HIV IN and achieved a sensitivity of 83nM. Since most pathogens encapsulate or encode their own suite of enzymes with unique activities, the developed CasPART method holds potential for broader applications in infectious disease detection and phenotypic characterization.

## Materials and Methods

### • In-house RNA synthesis

DNA templates were ordered via IDT gBlocks and amplified with PCR using primers with added T7 promoter sequence, followed by purification with PureLink PCR Purification kits (Invitrogen, Cat. No.: K310001). RNA was generated from the purified DNA templates using T7 RNA polymerase (NEB, Cat. No.: M0251L) following the manufacturer’s protocol. After incubating at 37°C for an hour, the RNA was purified using Monarch RNA Cleanup kits (NEB, Cat. No.: T2040L). To remove DNA templates in the RNA stock, purified RNA went through three rounds of DNA digestion using DNase I kits (NEB, Cat. No.: M0303L). Specifically, in each round, purified RNA was mixed with 10μL DNase I, 10μL 10X buffer included in the kit, 1μL RNasin Plus (Promega, Cat. No.: N2615), and water for a total of 100μL. After incubating at 37°C for an hour, the RNA was purified using the Monarch kit. The final RNA stock was quantified using the Qubit RNA High Sensitivity kit (Invitrogen, Cat. No.: Q32852). The length and integrity of the RNA were checked using Agilent High Sensitivity RNA ScreenTape (Agilent, Cat. No.: 5067-5579).

### • Quantify cDNA generation by HIV RT enzyme with qPCR

Recombinant HIV RT enzyme was ordered from Millipore Sigma (Cat. No.: 382129-500U). The mass concentration of the stock was converted to molar concentrations based on a molecular weight of 117kDa provided by the manufacturer. The 10μL reverse transcription reaction contained 1X HIV RT buffer (50mM Tris-HCl at pH 8, 50mM KCl, 10mM MgCl_2_, 2mM DTT, and 0.06% Triton X-100), 3mM DTT, 0.1% Triton X-100, 0.2U/μL RNasin Plus (Promega, Cat. No.: N2615), 0.4mM dNTPs, 200nM primer, and RNA templates. The reaction was incubated at 37°C followed by 10min at 95°C for HIV RT inactivation and RNA degradation. The reaction was then diluted 10-fold in water and 2μL diluted samples were added to 25μL qPCR reactions for cDNA quantification. The qPCR reactions contained 0.625U OneTaq Hot Start DNA polymerase (NEB, Cat. No.: M0481L), 1X OneTaq standard reaction buffer, 0.2mM dNTPs, 200nM forward primer, 200nM reverse primer, and 1X EvaGreen Dye (Biotium, Cat. No.: 31000). After an initial 2-minute heat spike at 95°C, the qPCR procedure includes 40 cycles of 15sec at 95°C, 15sec at 55°C and 30sec at 68°C. All sequence information can be found in the Supplementary Sequence Table.

### • LamPART assay with real-time fluorescence detection

A 10μL LamPART reaction contained 1X HIV RT buffer, 3mM DTT, 0.1% Triton X-100, 0.2U/μL RNasin Plus, 1.4mM dNTPs, 800nM FIP and BIP, 400nM LB and LF, 160nM B3, 300nM UDP quencher, 200nM UDP fluorophore, 400nM LB docker, 160nM B3 docker, 500nM TQ21 aptamer, 50nM RNA substrates, 26.25μg/mL TFpol. The master mix was incubated at 65°C for 5min and cooled down to room temperature before adding RNA substrates and TFpol, to allow proper hybridization of the dockers to primers and the folding of the aptamer. After HIV RT enzyme was added, the reaction was incubated at 37°C for 30min followed by 65°C for 1hr. All sequence information can be found in the Supplementary Sequence Table.

### • LamPART assay with endpoint LFA detection

The 10μL LamPART reaction was prepared the same as above except that no UDP quenchers or fluorophores were used and biotin-labeled FIP and FITC-labeled BIP were used. The reaction was incubated at 37°C for 30min followed by 65°C for 30min. The amplicons were detected using PCRD Flex Lateral Flow Strips (Abingdon Health, Cat. No.: FG-FD51676) following the manufacturer’s protocol. Specifically, the 10μL post-amplification reaction was added to 140μL extraction buffers provided in the kit. The lateral flow strip was then dipped inside the buffer for 10min. The strips were scanned using an Epson V700 Photo Scanner.

### • Immunocapture of HIV RT enzyme

250μg M-280 Sheep Anti-Mouse IgG Dynabeads (ThermoFisher, Cat. No.: 11201D) were washed three times with 500μL binding buffer (1X TBS at pH=7.4, 1% BSA) and incubated in 50μL binding buffer with 7.5μg mouse anti-HIV-RT IgG (MyBioSource, Cat. No.: MBS531805) at 4°C with rotation overnight. The beads were then washed three times with 500μL binding buffer with 0.05% Tween-20 and resuspended in 25μL lysis buffer (2X HIV RT buffer, 2% BSA, 1.2% NP-40). Pooled HIV-negative human plasma was obtained from BioIVT (BioIVT, Cat. No.: HUMANPLK2-0000291). Since the plasma sample was from a commercial vendor, our study was exempt from IRB requirements. 25μL pooled HIV-negative plasma was mixed with 25μL lysis buffer and spiked with different copy numbers of HIV RT enzyme and 1uL beads. Samples were then transferred into 0.2mL PCR tube strips (USA Scientific, Cat. No.: 1402-4700) and incubated at room temperature with rotation for 30min for HIV RT capture. After capture, the beads were washed three times with 200μL wash buffer (1X RT buffer, 0.1% BSA, 0.6% NP-40) using multi-channel pipettes and strip-tube magnetic racks (NEB, Cat. No.: S1515S).

For experiments quantifying capture efficiency with qPCR, 10μL reverse transcription reaction was added to the washed beads, followed by the same steps described in the section of quantifying cDNA generation activity of HIV RT enzyme by qPCR. The results were compared with freshly diluted HIV RT enzyme directly added to the reverse transcription reaction. For experiments demonstrating integrated workflow with LamPART, the washed beads was mixed 10μL LamPART reaction as described in the section of LamPART with real-time fluorescence detection or LamPART with endpoint LFA detection.

### • CasPART assay

100nM LbCas12a (NEB, Cat No.: M0653T) and 100nM crRNA was incubated in 1X HIV RT buffer at 25°C for 15min to form the Cas12a-crRNA complex. 10μL Cas12a-crRNA complex was then mixed with 10μL HIV RT reaction. The final 20μL reaction contained 1X HIV RT buffer, 5mM MgCl_2_, 5mM DTT, 0.4mM dNTPs, 500nM reporters, 0.2U/μL RNasin Plus, 50nM RT substrate, 200nM RT primer, and 50nM of the Cas12a-crRNA complex. The reaction was incubated at 37°C with fluorescence measurements every 2min. For experiments testing CasPART integrated with immunocapture, HIV RT enzyme was spiked into 25μL HIV-negative plasma and immunocaptured as described in the section of immunocapture of HIV RT enzyme. After capture, 20μL CasPART reaction described above was added to the wash beads followed by incubation at 37°C with fluorescence measurements every 2min. All sequence information can be found in the Supplementary Sequence Table.

### • Fitting rates from the real-time fluorescence curves in CasPART

In CRISPR-Cas nucleic acid detection systems, for a given copy number of the target strand, fluorescence generation exhibits a linear relationship with incubation time. In CasPART, the copy number of the target strand generated by HIV RT enzyme is linearly increasing with incubation time. Therefore, the relationship between fluorescence intensity and incubation time should follow a second order polynomial equation, *fluorescence* = *B*2 ∗ *t*^2^ + *B*1 ∗ *t* + *B*0, where B2 is associated with the rate of HIV RT enzyme generating the target strand. We fitted the real-time curves in CasPART to a second order polynomial equation and used B2 to represent the fitted rate. To balance the number of data points for fitting and avoiding the real-time curves deviating from the equation due to reagent depletion, we manually selected an RFU threshold of 4000 and used data points before the threshold for fitting the curves. All fitting was conducted in GraphPad Prism 10.

### • CasPART assay for measuring inhibition by rilpivirine

The CasPART assay was prepared as described above and included 1e7 copies of HIV RT enzyme. Rilpivirine (Fisher Scientific, Cat. No.: 50-194-8056) was diluted in DMSO, and 2μL was added to the reaction to limit DMSO concentrations in the final reaction. The reaction was incubated at 37°C for 2hr with fluorescence measurements every 2min. The fitted rates were obtained from the real-time curves as described above. Fitted rates from test samples with rilpivirine were normalized against the average of the control samples (no rilpivirine). The normalized signals were fitted to four-parameter logistic regression curves using GraphPad Prism 10.

### • CasPART for measuring inhibition of WT and MUT HIV RT enzyme to 3TC-TP

The CasPART assay was prepared in the same manner except 10μM dCTP was used (the concentrations of dATP, dGTP, and dTTP were kept the same at 0.4mM). The lamivudine triphosphate (3TC-TP) was ordered from Sierra Bioresearch. The mutant HIV RT enzyme was kindly provided by the NIH HIV Reagent Program (Cat. No.: ARP-3195).

For the inhibition responses across different concentrations of 3TC-TP, 1e7 copies of HIV RT enzyme, either WT or MUT, were included in CasPART. Fitted rates from test samples with 3TC-TP were normalized against the average of the control samples (no 3TC-TP). The normalized signals were fitted to four-parameter logistic regression curves using GraphPad Prism 10. For the inhibition responses across different copy numbers of HIV RT enzyme, 100μM 3TC-TP was included in CasPART. All reactions were incubated at 37°C for 2hr with fluorescence measurements every 2min.

### • CasPART adaption for HIV IN detection

To allow proper hybridization of the oligos into the substrate structure, 1μM of TS-5, TS-3, TS-3-Tail, and NTS was mixed in 1X HIV RT buffer and cooled down from 95°C to 4°C at a rate of 1°C per 3min. The prepared substrate was then stored in the fridge until use. HIV integrase was kindly provided by the NIH HIV Reagent Program (Cat. No.: HRP-20203). The assay was prepared the same as described in CasPART including the reaction components and concentrations, except that 50nM disintegration substrates were used instead of the RT substrate and RT primer. The reaction was incubated at 37°C with fluorescence measurements every 2min. All sequence information can be found in the Supplementary Sequence Table.

## Data Availability

All data produced in the present study are available upon reasonable request to the authors

## Acknowledgment

This work was supported by NIH/NIAID grants R33AI140460. We thank Bob Atkinson for his initial design in sequence #2 and the accompanying LAMP assay. We thank Joe Henthorn for his design in sequence #4 and the accompanying LAMP assay. We thank Cara Brainerd and Cosette Craig for sharing the materials and protocols of testing rilpivirine. We thank Dr. Joanne D. Stekler and Dr. Paul K. Drain for their clinical advice on the project. We thank Dr. Paul Yager, Dr. James I. Mullins, Dr. Patrick Stayton, and Dr. Xiaohu Gao for their advice on the project. Research reported in this publication was supported by the University of Washington/Fred Hutch Center for AIDS Research, an NIH-funded program under award number AI027757 which is supported by the following NIH Institutes and Centers: NIAID, NCI, NIMH, NIDA, NICHD, NHLBI, NIA, NIGMS, NIDDK. The content is solely the responsibility of the authors and does not necessarily represent the official views of the National Institutes of Health. The following reagents were obtained through the NIH HIV Reagent Program, Division of AIDS, NIAID, NIH: Human immunodeficiency virus type 1 HXB2 reverse transcriptase/MI84V heterodimeric protein, recombinant from Escherichia coli, ARP-3195, contributed by Dr. Vinayaka Prasad and Dr. Mark Wainberg; Integrase (F185K/C280S) protein from human immunodeficiency virus type 1 (HIV-1) NL4-3, recombinant from Escherichia coli, HRP-20203, contributed by DAIDS, NIAID. Schematics in the manuscript were created with BioRender.com.

## References

[1] A. Robinson, R.J. Causer, N.E. Dixon, Architecture and conservation of the bacterial DNA replication machinery, an underexploited drug target, Curr Drug Targets 13 (2012) 352–372.

[2] E.O. Freed, HIV-1 Replication, Somat Cell Mol Genet 26 (2001).

[3] H. Yang, Z. Rao, Structural biology of SARS-CoV-2 and implications for therapeutic development, Nat Rev Microbiol 19 (2021) 685–700. 10.1038/s41579-021-00630-8.

[4] T.K.H. Scheel, C.M. Rice, Understanding the hepatitis C virus life cycle paves the way for highly effective therapies, Nat Med 19 (2013) 837–849. 10.1038/nm.3248.

[5] E. De Clercq, G. Li, Approved antiviral drugs over the past 50 years, Clin Microbiol Rev 29 (2016) 695–747. 10.1128/CMR.00102-15.

[6] M.A. Kohanski, D.J. Dwyer, J.J. Collins, How antibiotics kill bacteria: From targets to networks, Nat Rev Microbiol 8 (2010) 423–435. 10.1038/nrmicro2333.

[7] C.J. Murray, K.S. Ikuta, F. Sharara, L. Swetschinski, G. Robles Aguilar, A. Gray, C. Han, C. Bisignano, P. Rao, E. Wool, S.C. Johnson, A.J. Browne, M.G. Chipeta, F. Fell, S. Hackett, G. Haines-Woodhouse, B.H. Kashef Hamadani, E.A.P. Kumaran, B. McManigal, R. Agarwal, S. Akech, S. Albertson, J. Amuasi, J. Andrews, A. Aravkin, E. Ashley, F. Bailey, S. Baker, B. Basnyat, A. Bekker, R. Bender, A. Bethou, J. Bielicki, S. Boonkasidecha, J. Bukosia, C. Carvalheiro, C. Castañeda-Orjuela, V. Chansamouth, S. Chaurasia, S. Chiurchiù, F. Chowdhury, A.J. Cook, B. Cooper, T.R. Cressey, E. Criollo-Mora, M. Cunningham, S. Darboe, N.P.J. Day, M. De Luca, K. Dokova, A. Dramowski, S.J. Dunachie, T. Eckmanns, D. Eibach, A. Emami, N. Feasey, N. Fisher-Pearson, K. Forrest, D. Garrett, P. Gastmeier, A.Z. Giref, R.C. Greer, V. Gupta, S. Haller, A. Haselbeck, S.I. Hay, M. Holm, S. Hopkins, K.C. Iregbu, J. Jacobs, D. Jarovsky, F. Javanmardi, M. Khorana, N. Kissoon, E. Kobeissi, T. Kostyanev, F. Krapp, R. Krumkamp, A. Kumar, H.H. Kyu, C. Lim, D. Limmathurotsakul, M.J. Loftus, M. Lunn, J. Ma, N. Mturi, T. Munera-Huertas, P. Musicha, M.M. Mussi-Pinhata, T. Nakamura, R. Nanavati, S. Nangia, P. Newton, C. Ngoun, A. Novotney, D. Nwakanma, C.W. Obiero, A. Olivas-Martinez, P. Olliaro, E. Ooko, E. Ortiz-Brizuela, A.Y. Peleg, C. Perrone, N. Plakkal, A. Ponce-de-Leon, M. Raad, T. Ramdin, A. Riddell, T. Roberts, J.V. Robotham, A. Roca, K.E. Rudd, N. Russell, J. Schnall, J.A.G. Scott, M. Shivamallappa, J. Sifuentes-Osornio, N. Steenkeste, A.J. Stewardson, T. Stoeva, N. Tasak, A. Thaiprakong, G. Thwaites, C. Turner, P. Turner, H.R. van Doorn, S. Velaphi, A. Vongpradith, H. Vu, T. Walsh, S. Waner, T. Wangrangsimakul, T. Wozniak, P. Zheng, B. Sartorius, A.D. Lopez, A. Stergachis, C. Moore, C. Dolecek, M. Naghavi, Global burden of bacterial antimicrobial resistance in 2019: a systematic analysis, The Lancet 399 (2022) 629–655. 10.1016/S0140-6736(21)02724-0.

[8] M.E. Cilento, K.A. Kirby, S.G. Sarafianos, Avoiding drug resistance in HIV reverse transcriptase, Chem Rev 121 (2021) 3271–3296. 10.1021/acs.chemrev.0c00967.

[9] S.Y. Rhee, P.M. Grant, P.L. Tzou, G. Barrow, P.R. Harrigan, J.P.A. Ioannidis, R.W. Shafer, A systematic review of the genetic mechanisms of dolutegravir resistance, Journal of Antimicrobial Chemotherapy 74 (2019) 3135–3149. 10.1093/jac/dkz256.

[10] T. Shaw, A. Bartholomeusz, S. Locarnini, HBV drug resistance: Mechanisms, detection and interpretation, J Hepatol 44 (2006) 593–606. 10.1016/j.jhep.2006.01.001.

[11] C. Sarrazin, J.W. Goethe, The importance of resistance to direct antiviral drugs in HCV infection in clinical practice, J Hepatol 64 (2016) 486–504.

[12] S. Gandhi, J. Klein, A.J. Robertson, M.A. Peña-Hernández, M.J. Lin, P. Roychoudhury, P. Lu, J. Fournier, D. Ferguson, S.A.K. Mohamed Bakhash, M. Catherine Muenker, A. Srivathsan, E.A. Wunder, N. Kerantzas, W. Wang, B. Lindenbach, A. Pyle, C.B. Wilen, O. Ogbuagu, A.L. Greninger, A. Iwasaki, W.L. Schulz, A.I. Ko, De novo emergence of a remdesivir resistance mutation during treatment of persistent SARS-CoV-2 infection in an immunocompromised patient: a case report, Nat Commun 13 (2022). 10.1038/s41467-022-29104-y.

[13] D.H. Goldhill, A.J.W. Te Velthuis, R.A. Fletcher, P. Langat, M. Zambon, A. Lackenby, W.S. Barclay, The mechanism of resistance to favipiravir in influenza, Proc Natl Acad Sci U S A 115 (2018) 11613–11618. 10.1073/pnas.1811345115.

[14] F.G. Hayden, N. Sugaya, N. Hirotsu, N. Lee, M.D. de Jong, A.C. Hurt, T. Ishida, H. Sekino, K. Yamada, S. Portsmouth, K. Kawaguchi, T. Shishido, M. Arai, K. Tsuchiya, T. Uehara, A. Watanabe, Baloxavir marboxil for uncomplicated influenza in adults and adolescents, New England Journal of Medicine 379 (2018) 913–923. 10.1056/nejmoa1716197.

[15] J. Piret, G. Boivin, Resistance of herpes simplex viruses to nucleoside analogues: Mechanisms, prevalence, and management, Antimicrob Agents Chemother 55 (2011) 459–472. 10.1128/AAC.00615-10.

[16] B.P. Goldstein, Resistance to rifampicin: A review, Journal of Antibiotics 67 (2014) 625–630. 10.1038/ja.2014.107.

[17] V. Tchesnokova, L. Larson, I. Basova, Y. Sledneva, D. Choudhury, T. Solyanik, J. Heng, T.C. Bonilla, S. Pham, E.M. Schartz, L.T. Madziwa, E. Holden, S.J. Weissman, J.D. Ralston, E. V. Sokurenko, Increase in the community circulation of ciprofloxacin-resistant Escherichia coli despite reduction in antibiotic prescriptions, Communications Medicine 3 (2023). 10.1038/s43856-023-00337-2.

[18] D.C. Hooper, G.A. Jacoby, Mechanisms of drug resistance: Quinolone resistance, Ann N Y Acad Sci 1354 (2015) 12–31. 10.1111/nyas.12830.

[19] World Health Organization, Consolidated guidelines on HIV prevention, testing, treatment, service delivery and monitoring: recommendations for a public health approach., (n.d.).

[20] B.S. Parekh, C.Y. Ou, P.N. Fonjungo, M.B. Kalou, E. Rottinghaus, A. Puren, H. Alexander, M.H. Cox, J.N. Nkengasong, Diagnosis of human immunodeficiency virus infection, Clin Microbiol Rev 32 (2019). 10.1128/CMR.00064-18.

[21] K.J. Metzner, Technologies for HIV-1 drug resistance testing: inventory and needs, Curr Opin HIV AIDS 17 (2022) 222–228. 10.1097/COH.0000000000000737.

[22] N.B.Y. Tsui, E.K.O. Ng, Y.M. Dennis Lo, Stability of endogenous and added RNA in blood specimens, serum, and plasma, Clin Chem 48 (2002) 1647–1653. https://academic.oup.com/clinchem/article/48/10/1647/5642225.

[23] J. Hemelaar, The origin and diversity of the HIV-1 pandemic, Trends Mol Med 18 (2012) 182–192. 10.1016/j.molmed.2011.12.001.

[24] K.A. Curtis, D. Morrison, D.L. Rudolph, A. Shankar, L.S.P. Bloomfield, W.M. Switzer, S.M. Owen, A multiplexed RT-LAMP assay for detection of group M HIV-1 in plasma or whole blood, J Virol Methods 255 (2018) 91–97. 10.1016/j.jviromet.2018.02.012.

[25] W.-S. Hu, S.H. Hughes, HIV-1 reverse transcription, Cold Spring Harbor Perspective in Medicine 2 (2012). 10.1101/cshperspect.a006882.

[26] W. Heneine, S. Yamamoto, W.M. Switzer, T.J. Spira, T.M. Folks, Detection of reverse transcriptase by a highly sensitive assay in sera from persons infected with human immunodeficiency virus type 1, J Infect Dis 171 (1995) 1210–1216.

27. H. Pyra, J. Boni, J. Schupbach, Ultrasensitive retrovirus detection by a reverse transcriptase assay based on product enhancement, Proc. Natl. Acad. Sci. USA 91 (1994) 1544–1548.

[28] A. Malmsten, X.-W. Shao, S. Sjödahl, E.-L. Fredriksson, I. Pettersson, T. Leitner, C.F.R. Källander, E. Sandström, J.S. Gronowitz, Improved HIV-1 viral load determination based on reverse transcriptase activity recovered from human plasma, J Med Virol 76 (2005) 291–296. 10.1002/jmv.20360.

[29] P. Burgisser, P. Vernazza, M. Flepp, J. Boni, Z. Tomasik, U. Hummel, G. Pantaleo, J. Schupbach, Swiss HIV Cohort Study, Performance of five different assays for the quantification of viral load in persons infected with various subtypes of HIV-1, J Acquir Immune Defic Syndr (1988) 23 (2000) 138–144.

[30] X.-W. Shao, A. Malmsten, J. Lennerstrand, A. Sönnerborg D, T. Unge, S.J. Gronowitz, C.F.R. Källander, Use of HIV-1 reverse transcriptase recovered from human plasma for phenotypic drug susceptibility testing, AIDS 17 (2003) 1463–1471. 10.1097/01.aids.0000072670.21517.0d.

[31] D. Hoffmann, A.D. Garcia, P.R. Harrigan, I.C.D. Johnston, T. Nakasone, J.G. García-Lerma, W. Heneine, Measuring enzymatic HIV-1 susceptibility to two reverse transcriptase inhibitors as a rapid and simple approach to HIV-1 drug-resistance testing, PLoS One 6 (2011). 10.1371/journal.pone.0022019.

[32] S.P. Layne, M.J. Merges, M. Dembo, J.L. Spouge, S.R. Conley, J.P. Moore, J.L. Raina, H. Renz, H.R. Gelderblom, P.L. Nara, Factors underlying spontaneous inactivation and susceptibility to neutralization of human immunodeficiency virus, Virology 189 (1992) 695–714.

[33] F. Marino-Merlo, B. Macchi, D. Armenia, M.C. Bellocchi, F. Ceccherini-Silberstein, A. Mastino, S. Grelli, Focus on recently developed assays for detection of resistance/sensitivity to reverse transcriptase inhibitors, Appl Microbiol Biotechnol (2018) 9925–9936. 10.1007/s00253-018-9390-x.

[34] T. Notomi, H. Okayama, H. Masubuchi, T. Yonekawa, K. Watanabe, N. Amino, T. Hase, Loop-mediated isothermal amplification of DNA, Nucleic Acids Res 28 (2000) 63.

[35] N. Panpradist, E.C. Kline, R.G. Atkinson, M. Roller, Q. Wang, I.T. Hull, J.H. Kotnik, A.K. Oreskovic, C. Bennett, D. Leon, V. Lyon, S.D. Gilligan-Steinberg, P.D. Han, P.K. Drain, L.M. Starita, M.J. Thompson, B.R. Lutz, Harmony COVID-19: A ready-to-use kit, low-cost detector, and smartphone app for point-of-care SARS-CoV-2 RNA detection, Sci Adv 7 (2021) 1281. https://www.science.org.

[36] C. Myhrvold, C.A. Freije, J.S. Gootenberg, O.O. Abudayyeh, H.C. Metsky, A.F. Durbin, M.J. Kellner, A.L. Tan, L.M. Paul, L.A. Parham, K.F. Garcia, K.G. Barnes, B. Chak, A. Mondini, M.L. Nogueira, S. Isern, S.F. Michael, I. Lorenzana, N.L. Yozwiak, B.L. MacInnis, I. Bosch, L. Gehrke, F. Zhang, P.C. Sabeti, Field-deployable viral diagnostics using CRISPR-Cas13, Science (1979) 360 (2018) 444–448. https://www.science.org.

[37] J.S. Chen, E. Ma, L.B. Harrington, M. Da Costa, X. Tian, J.M. Palefsky, J.A. Doudna, CRISPR-Cas12a target binding unleashes indiscriminate single-stranded DNase activity, Science (1979) 360 (2018) 436–439.

[38] M.M. Kaminski, O.O. Abudayyeh, J.S. Gootenberg, F. Zhang, J.J. Collins, CRISPR-based diagnostics, Nat Biomed Eng 5 (2021) 643–656. 10.1038/s41551-021-00760-7.

[39] O.O. Abudayyeh, J.S. Gootenberg, CRISPR diagnostics, Science (1979) 372 (2021) 914–915.

[40] S. Yamamoto, T.M. Folks, W. Heneine, Highly sensitive qualitative and quantitative detection of reverse transcriptase activity: optimization, validation, and comparative analysis with other detection systems, J Virol Methods 61 (1996) 135–143. 10.1016/0166-0934(96)02078-2.

[41] M.D. Jones, N.S. Foulkes, Reverse transcription of mRNA by Thermus aquaticus DNA polymerase, Nucleic Acids Res 17 (1989) 8387–8388.

[42] G. Divita, U. Immendorfer, M. Gautel, K. Rittinger, T. Restle, R.S. Goody, Kinetics of interaction of HIV reverse transcriptase with primer/template, Biochemistry 32 (1993) 7966–7971. https://pubs.acs.org/sharingguidelines (accessed January 30, 2020).

[43] E.C. Kline, N. Panpradist, I.T. Hull, Q. Wang, A.K. Oreskovic, P.D. Han, L.M. Starita, B.R. Lutz, Multiplex target-redundant RT-LAMP for robust detection of SARS-CoV-2 using fluorescent universal displacement probes, Microbiol Spectr 10 (2022). 10.1128/spectrum.01583-21.

[44] C. Shi, X. Shen, S. Niu, C. Ma, Innate reverse transcriptase activity of DNA polymerase for isothermal RNA direct detection, J Am Chem Soc 137 (2015) 13804–13806. 10.1021/jacs.5b08144.

[45] C. Dang, S.D. Jayasena, Oligonucleotide inhibitors of Taq DNA polymerase facilitate detection of low copy number targets by PCR, J Mol Biol 264 (1996) 268–278. 10.1006/JMBI.1996.0640.

[46] Y. Lin, S.D. Jayasena, Inhibition of multiple thermostable DNA polymerases by a heterodimeric aptamer, J Mol Biol 271 (1997) 100–111. 10.1006/JMBI.1997.1165.

[47] T.L. Dangerfield, I. Paik, S. Bhadra, K.A. Johnson, A.D. Ellington, Kinetics of elementary steps in loop-mediated isothermal amplification (LAMP) show that strand invasion during initiation is rate-limiting, Nucleic Acids Res 51 (2023) 488–499. 10.1093/nar/gkac1221.

[48] Y. Chen, N. Cheng, Y. Xu, K. Huang, Y. Luo, W. Xu, Point-of-care and visual detection of P. aeruginosa and its toxin genes by multiple LAMP and lateral flow nucleic acid biosensor, Biosens Bioelectron 81 (2016) 317–323. 10.1016/j.bios.2016.03.006.

[49] A. Chang, J.M. Ostrove, R.E. Bird, Development of an improved product enhanced reverse transcriptase assay, J Virol Methods 65 (1997) 45–54. 10.1016/S0166-0934(96)02168-4.

[50] P. Mikel, P. Vasickova, R. Tesarik, H. Malenovska, P. Kulich, T. Vesely, P. Kralik, Preparation of MS2 phage-like particles and their use as potential process control viruses for detection and quantification of enteric RNA viruses in different matrices, Front Microbiol 7 (2016). 10.3389/fmicb.2016.01911.

[51] N. Panpradist, Q. Wang, P.S. Ruth, J.H. Kotnik, A.K. Oreskovic, A. Miller, S.W.A. Stewart, J. Vrana, P.D. Han, I.A. Beck, L.M. Starita, L.M. Frenkel, B.R. Lutz, Simpler and faster Covid-19 testing: strategies to streamline SARS-CoV-2 molecular assays, EBioMedicine 64 (2021). 10.1016/j.ebiom.2021.103236.

[52] L.T. Nguyen, B.M. Smith, P.K. Jain, Enhancement of trans-cleavage activity of Cas12a with engineered crRNA enables amplified nucleic acid detection, Nat Commun 11 (2020). 10.1038/s41467-020-18615-1.

[53] P. Fozouni, S. Son, M. Díaz de León Derby, G.J. Knott, C.N. Gray, M. V. D’Ambrosio, C. Zhao, N.A. Switz, G.R. Kumar, S.I. Stephens, D. Boehm, C.L. Tsou, J. Shu, A. Bhuiya, M. Armstrong, A.R. Harris, P.Y. Chen, J.M. Osterloh, A. Meyer-Franke, B. Joehnk, K. Walcott, A. Sil, C. Langelier, K.S. Pollard, E.D. Crawford, A.S. Puschnik, M. Phelps, A. Kistler, J.L. DeRisi, J.A. Doudna, D.A. Fletcher, M. Ott, Amplification-free detection of SARS-CoV-2 with CRISPR-Cas13a and mobile phone microscopy, Cell 184 (2021) 323–333. 10.1016/j.cell.2020.12.001.

[54] Y. Liu, L. Zhan, Z. Qin, J. Sackrison, J.C. Bischof, Ultrasensitive and highly specific lateral flow assays for point-of-care diagnosis, ACS Nano 15 (2021) 3593–3611. 10.1021/acsnano.0c10035.

[55] A.O. Olanrewaju, B.P. Sullivan, J.Y. Zhang, A.T. Bender, D. Sevenler, T.J. Lo, M. Fernandez-Suarez, P.K. Drain, J.D. Posner, Enzymatic assay for rapid measurement of antiretroviral drug levels, ACS Sens 5 (2020) 952–959. 10.1021/ACSSENSORS.9B02198/SUPPL_FILE/SE9B02198_SI_001.PDF.

[56] A.O. Olanrewaju, B.P. Sullivan, A.H. Gim, C.A. Craig, D. Sevenler, A.T. Bender, P.K. Drain, J.D. Posner, REverSe TRanscrIptase chain termination (RESTRICT) for selective measurement of nucleotide analogs used in HIV care and prevention, Bioeng Transl Med 8 (2023). 10.1002/btm2.10369.

[57] French National AIDS Agency, Recommendations for therapeutic drug monitoring of CABOTEGRAVIR and RILPIVIRINE during long-acting injectable administration of Vocabria/Rekambys every 2 months in HIV-infected patients, (2022).

[58] C.A.B. Boucher, N. Cammack, P. Schipper, R. Schuurman, P. Rouse, M.A. Wainberg, J.M. Cameron, High-Level resistance to (-) enantiomeric 2’-deoxy-3’-thiacytidine in vitro is due to one amino acid substitution in the catalytic site of human immunodeficiency virus type 1 reverse transcriptase, Antimicrob Agents Chemother 37 (1993) 2231–2234. https://journals.asm.org/journal/aac.

[59] J.G.G. Lerma, R.F. Schinazi, A.S. Juodawlkis, V. Soriano, Y. Lin, K. Tatti, D. Rimland, T.M. Folks, W. Heneine, A rapid non-culture-based assay for clinical monitoring of phenotypic resistance of human immunodeficiency virus type 1 to lamivudine (3TC), Antimicrob Agents Chemother 43 (1999) 264–270.

[60] J. Deval, K.L. White, M.D. Miller, N.T. Parkin, J. Courcambeck, P. Halfon, B. Selmi, J. Boretto, B. Canard, Mechanistic basis for reduced viral and enzymatic fitness of HIV-1 reverse transcriptase containing both K65R and M184V Mutations, J Biol Chem 279 (2004) 509–516. 10.1074/jbc.M308806200.

[61] C.A. Cannon, M.S. Ramchandani, M.R. Golden, Feasibility of a novel self-collection method for blood samples and its acceptability for future home-based PrEP monitoring, BMC Infect Dis 22 (2022). 10.1186/s12879-022-07432-0.

[62] G. Vázquez-Rosales, J.G.G. Lerma, S. Yamamoto, W.M. Switzer, D. Havlir, T.M. Folks, D.D. Richman, W. Heneine, Rapid screening of phenotypic resistance to nevirapine by direct analysis of HIV type 1 reverse transcriptase activity in plasma, AIDS Res Hum Retroviruses 15 (1999) 1191–1200. www.liebertpub.com (accessed June 1, 2021).

[63] T.L. Diamond, F.D. Bushman, Role of metal ions in catalysis by HIV integrase analyzed using a quantitative PCR disintegration assay, Nucleic Acids Res 34 (2006) 6116–6125. 10.1093/nar/gkl862.

[64] S.A. Chow, K.A. Vincent, V. Ellison, P.O. Brown, Reversal of integration and DNA splicing mediated by integrase of human immunodeficiency virus, Science (1979) 255 (1992) 723–726.

[65] R. Craigie, F.D. Bushman, HIV DNA integration, Cold Spring Harb Perspect Med 2 (2012).

[66] R. Craigie, The molecular biology of HIV integrase, Future Virol 7 (2012) 679–686. 10.2217/fvl.12.56.

