## Supplementary Information for "Sensitive Pathogen Detection and Drug Resistance Characterization Using Pathogen-Derived Enzyme Activity Amplified by LAMP or CRISPR-Cas"

**Table of Contents**

| **page S4** | Linear correlations of cDNA copy number versus HIV RT copy number within 30min or 1hr incubation.  **Figure S1.** |
| --- | --- |
| **page S5** | Linear correlations of cDNA copy number versus HIV RT copy number with different concentrations of RNA substrates.  **Figure S2.** |
| **page S6** | HIV RT efficiency on different sequences.  **Figure S3.** |
| **page S7** | Innate reverse transcription activity of Bst 2.0 WarmStart (WS) and TF Pol.  **Figure S4.** |
| **page S8** | The cDNA copy number per HIV RT versus RNA concentrations under LAMP conditions.  **Figure S5.** |
| **page S9** | Results of 20 replicates of one-pot LamPART with 10 copies of HIV RT.  **Figure S6.** |
| **page S10** | Illustrations of the endpoint lateral flow detection of LAMP amplicons.  **Figure S7.** |
| **page S11** | The cDNA copy number generated by free HIV RT versus HIV RT preincubated with antibodies.  **Figure S8.** |
| **page S12** | High-throughput testing with the integrated workflow by using strip tubes and multi-channel pipettes.  **Figure S9.** |
| **page S13** | CasPART signals using substrates with two activation regions versus a single activation region.  **Figure S10.** |
| **page S14** | Real-time curves of final CasPART performance during 5hr incubation.  **Figure S11.** |
| **page S15** | CasPART real-time curves and fitted rates of HIV RT enzyme with different concentrations of rilpivirine.  **Figure S12 and Supplementary Table S1.** |
| **page S16** | Concentrations of dCTP on CasPART signals.  **Figure S13** |
| **page S17** | CasPART real-time curves and fitted rates of wild-type and mutant HIV RT enzyme with different concentrations of 3TC-TP.  **Figure S14, Supplementary Table S2 and S3.** |
| **page S18-S19** | CasPART real-time curves and fitted rates of wild-type and mutant HIV RT enzyme at different copy numbers with or without 3TC-TP.  **Figure S15, Supplementary Table S4, S5, S6 and S7.** |
| **page S20** | PCR quantification of HIV integrase activity.  **Figure S16.** |
| **page S21-S23** | **Sequence table.** |

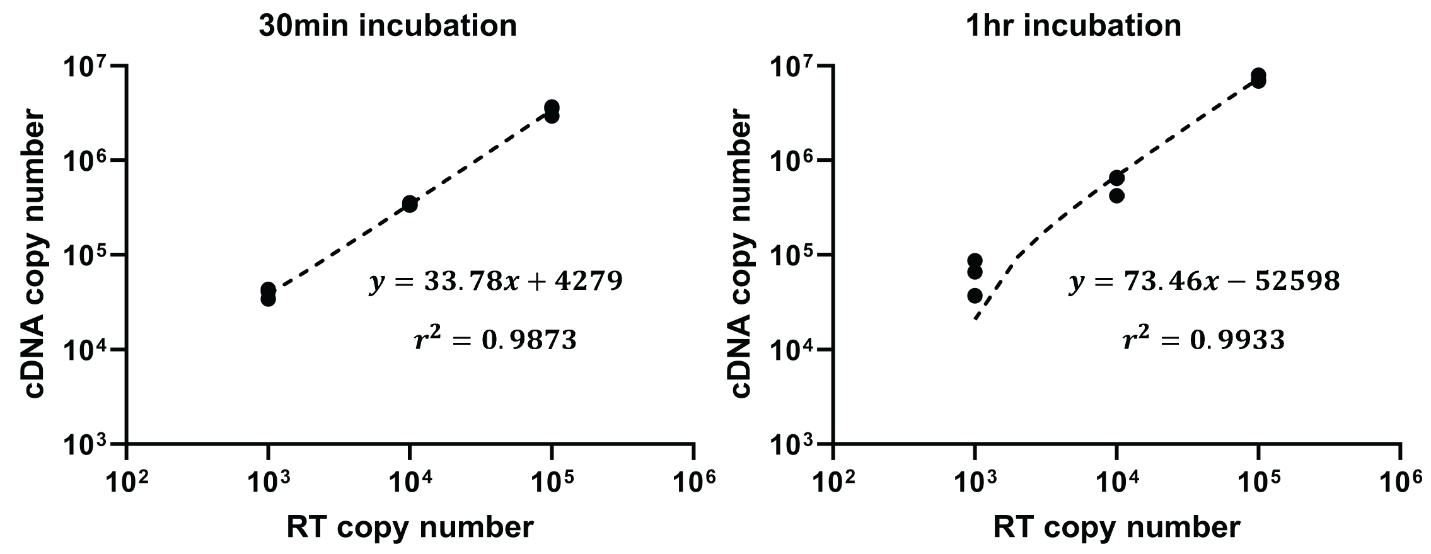

Figure S1 Linear correlations of cDNA copy number versus HIV RT copy number with 30min or 1hr incubation. 5nM RNA substrates were used.

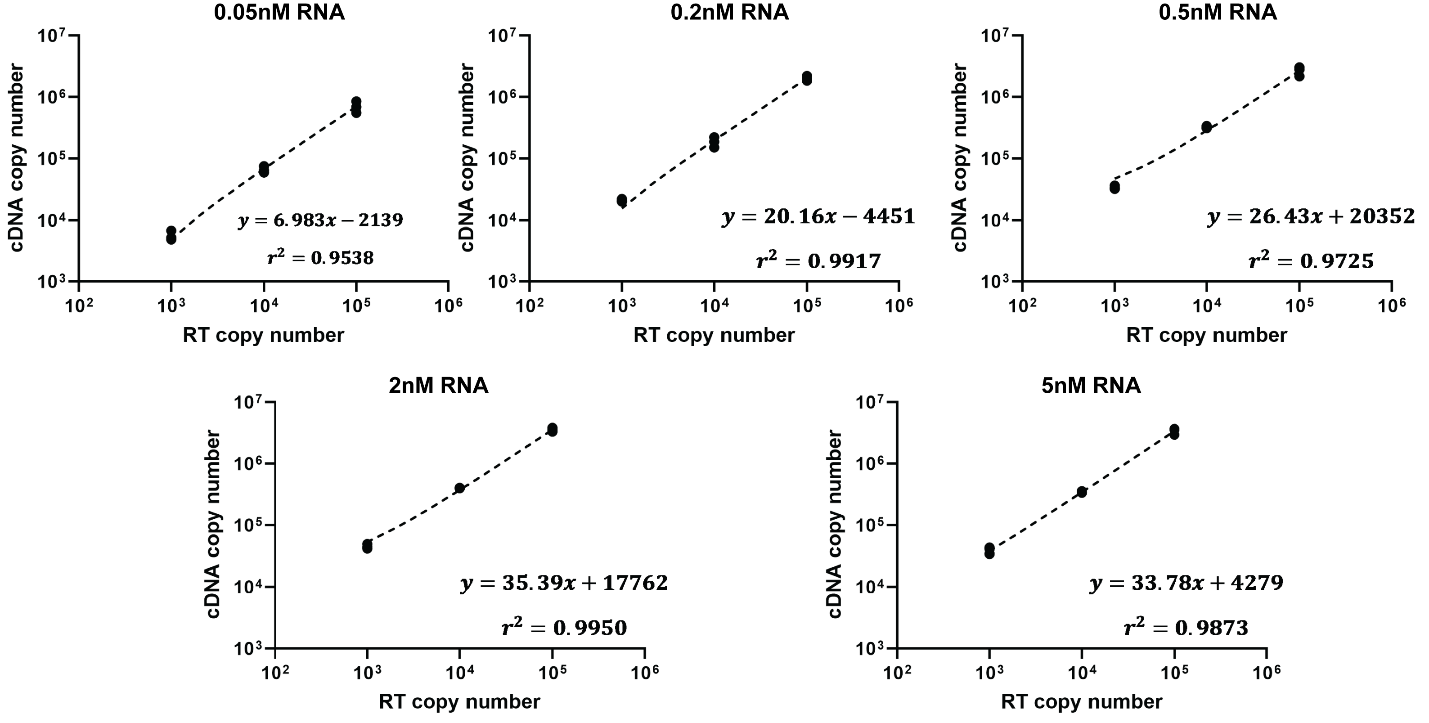

Figure S2 Linear correlations of cDNA copy number versus HIV RT copy number with different concentrations of RNA substrates. The RT reaction was incubated for 30min.

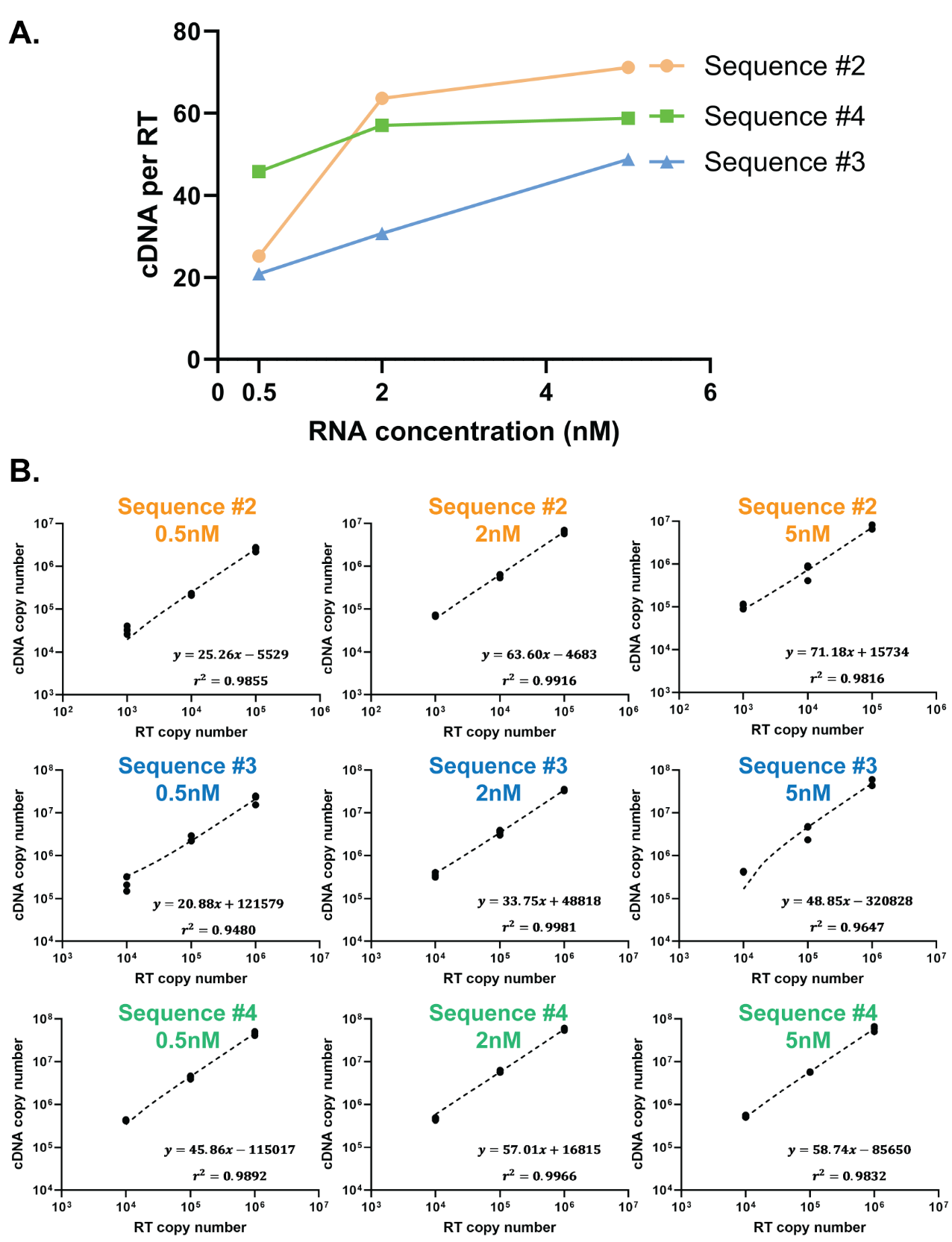

Figure S3 HIV RT efficiency on different sequences. (A) The cDNA per RT with 0.5nM, 2nM and 5nM of RNA sequence #2, #3 and #4. (B) The linear correlations of cDNA copy number against HIV RT copy number at each data point in panel A.

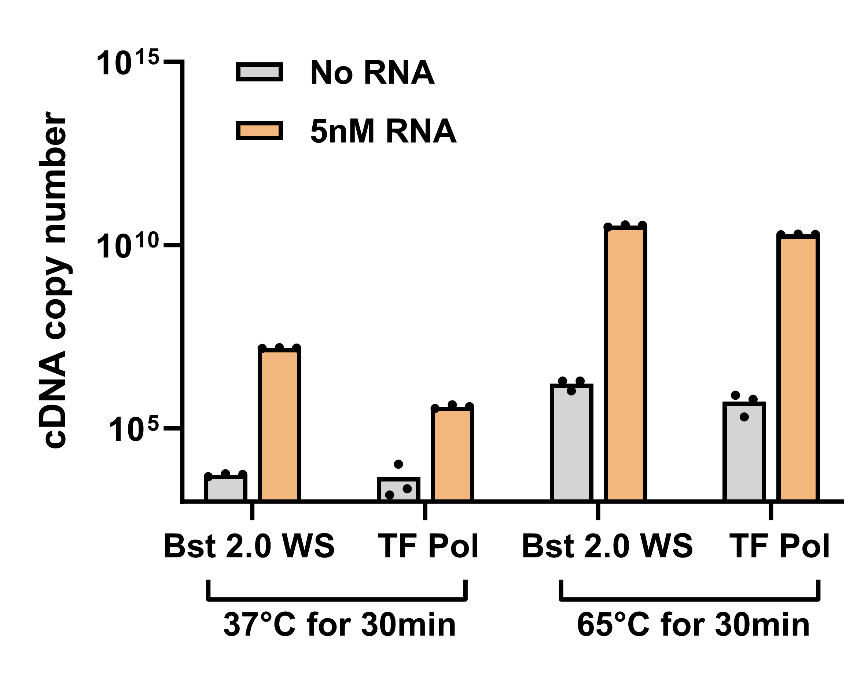

Figure S4 Innate reverse transcription activity of Bst 2.0 WarmStart (WS) and TF Pol. Bst 2.0 WS and TF Pol at concentrations used in LAMP were added to the same reverse transcription reaction used to quantify cDNA generation by HIV RT, with 5nM RNA sequence #2 as the substrate. After incubating at 37°C or 65°C for 30min, the reaction was heated at 95°C for 20min to inactivate Bst2.0 WS or TF Pol, followed by dilution and qPCR quantification, the same protocol used to quantify cDNA generation by HIV RT.

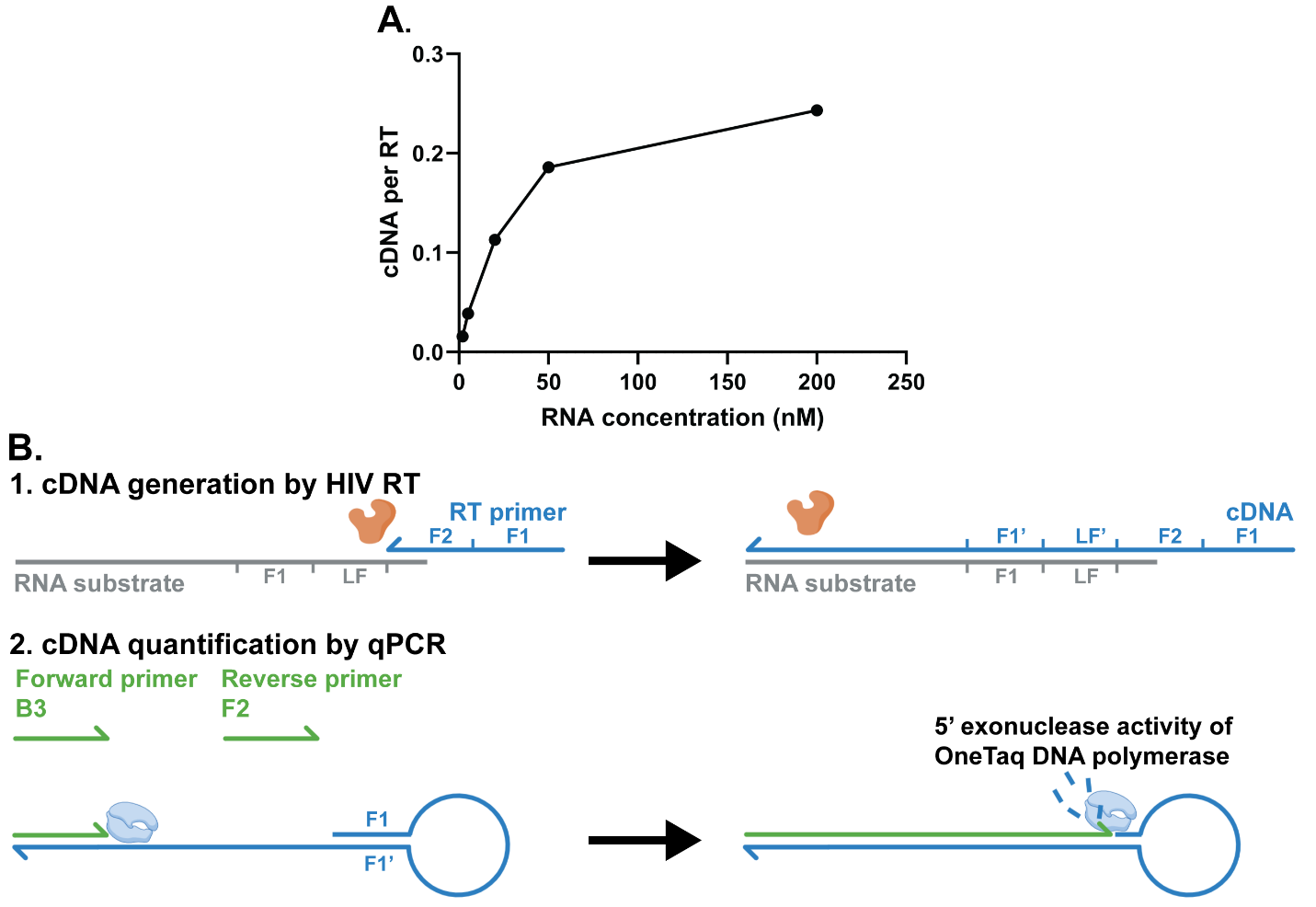

Figure S5 (A) The cDNA per HIV RT versus RNA concentrations under LAMP conditions, as quantified by qPCR. Note that the cDNA per HIV RT was much less compared to results in Figure 1, which could be due to the PCR quantification artifact illustrated in panel B. (B) Due to the truncated RNA substrate design and use of FIP (F1-F2) as the RT primer, the cDNA generated by HIV RT contains a hairpin loop structure on the 5’ end. While the cDNA is amplifiable in qPCR owing to the 5’ exonuclease activity of the OneTaq DNA polymerase in PCR, the cDNA template may have a lower amplification efficiency compared to the qPCR reference standards which do not have the hairpin structure, inducing an artifact of lower quantification outcomes. Nevertheless, the quantification artifact does not affect the overall trend of the curve in panel A.

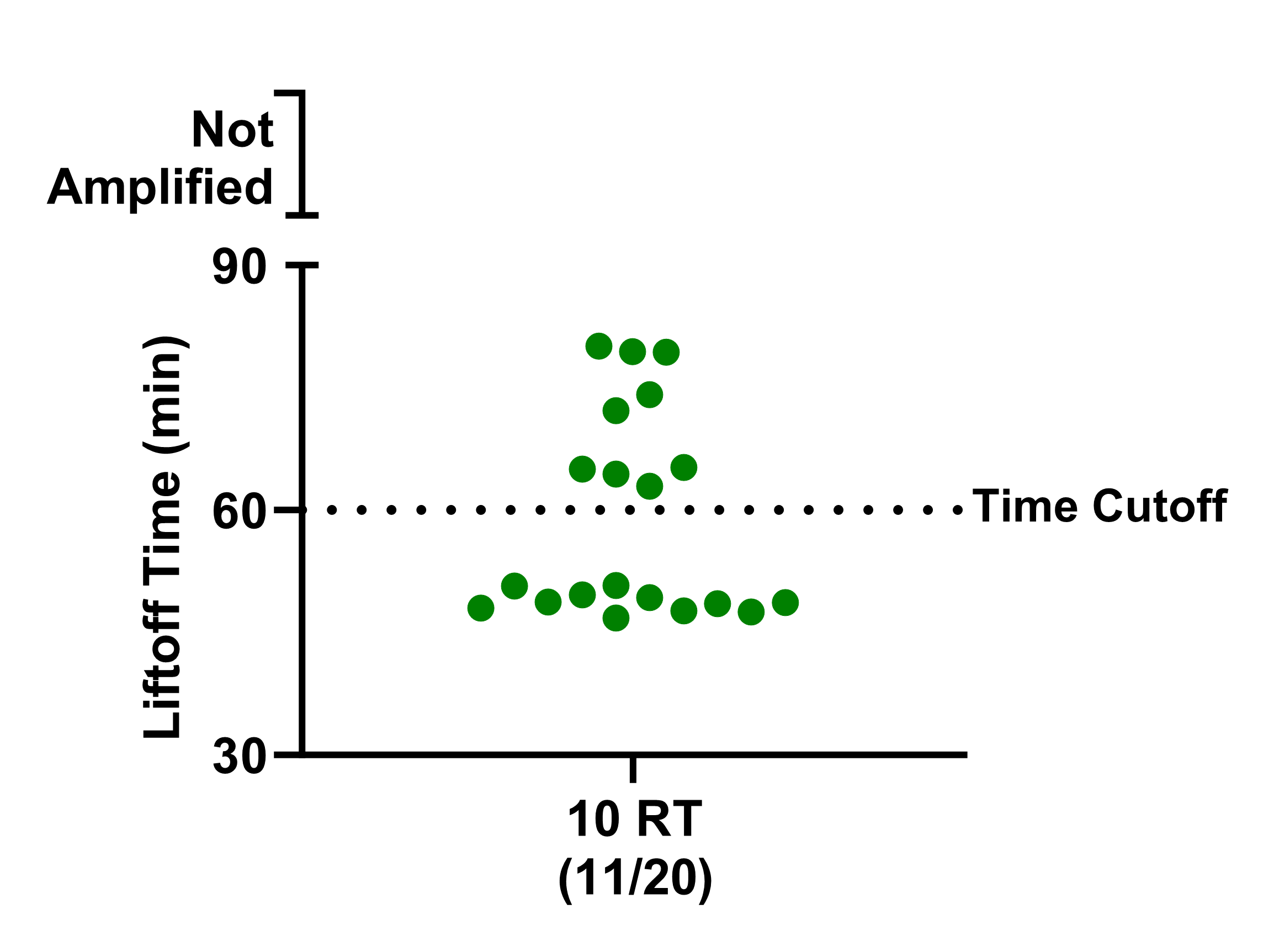

Figure S6 One-pot LamPART results of 20 replicates of 10-RT samples.

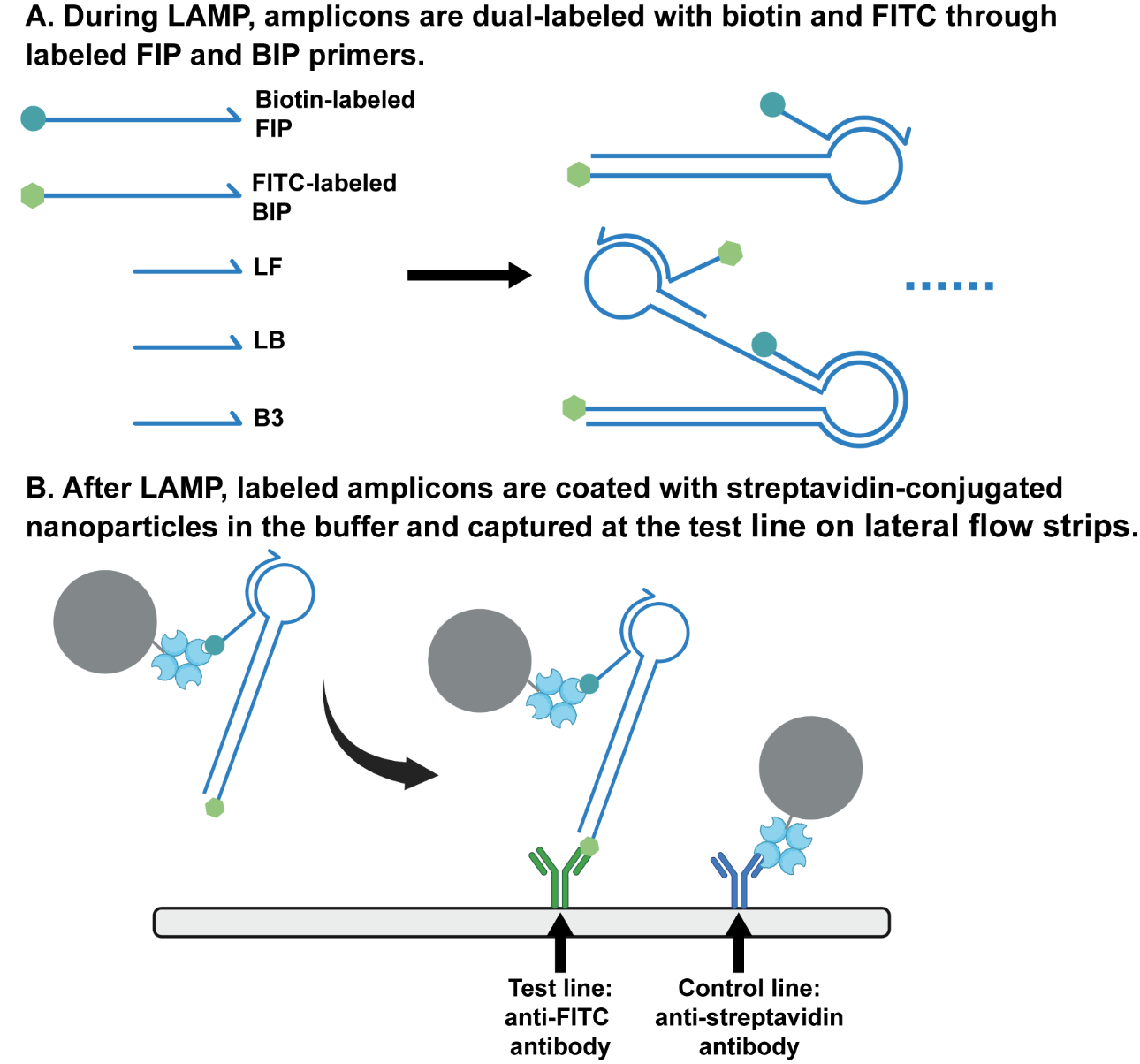

Figure S7 Illustrations of the endpoint lateral flow detection of LAMP amplicons.

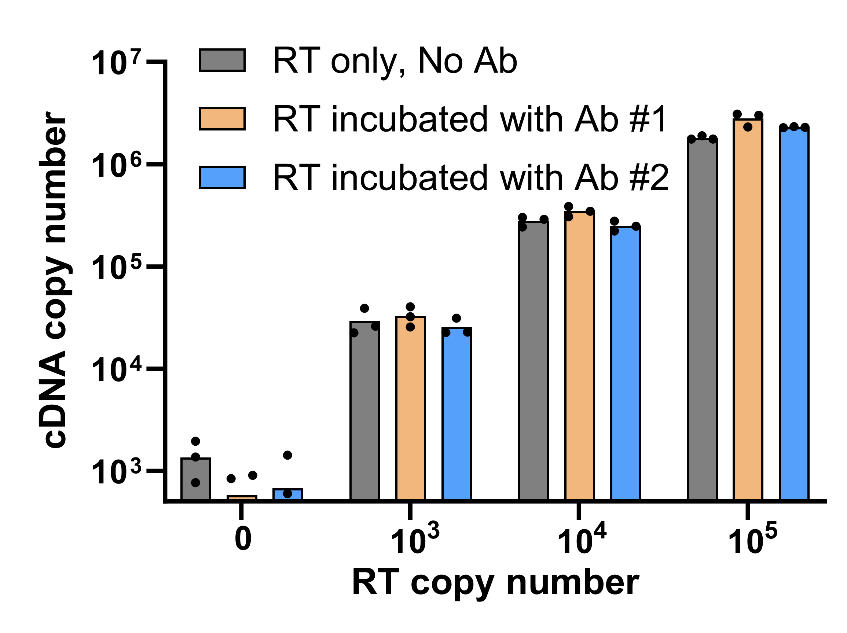

Figure S8 The cDNA copy number generated by free HIV RT versus HIV RT preincubated with antibodies. Free HIV RT or HIV RT mixed with 15μg/mL of antibody #1 (MyBioSource, Cat. No.: MBS531805) or antibody #2 (MyBioSource, Cat. No.: MBS601420) in buffer (1X TBS at pH=7.4, 1% BSA) was incubated in a rotator at room temperature for 30min, followed by quantifying cDNA generation by qPCR as described in HIV RT activity characterization. Ab #1 was selected to move forward for assay development.

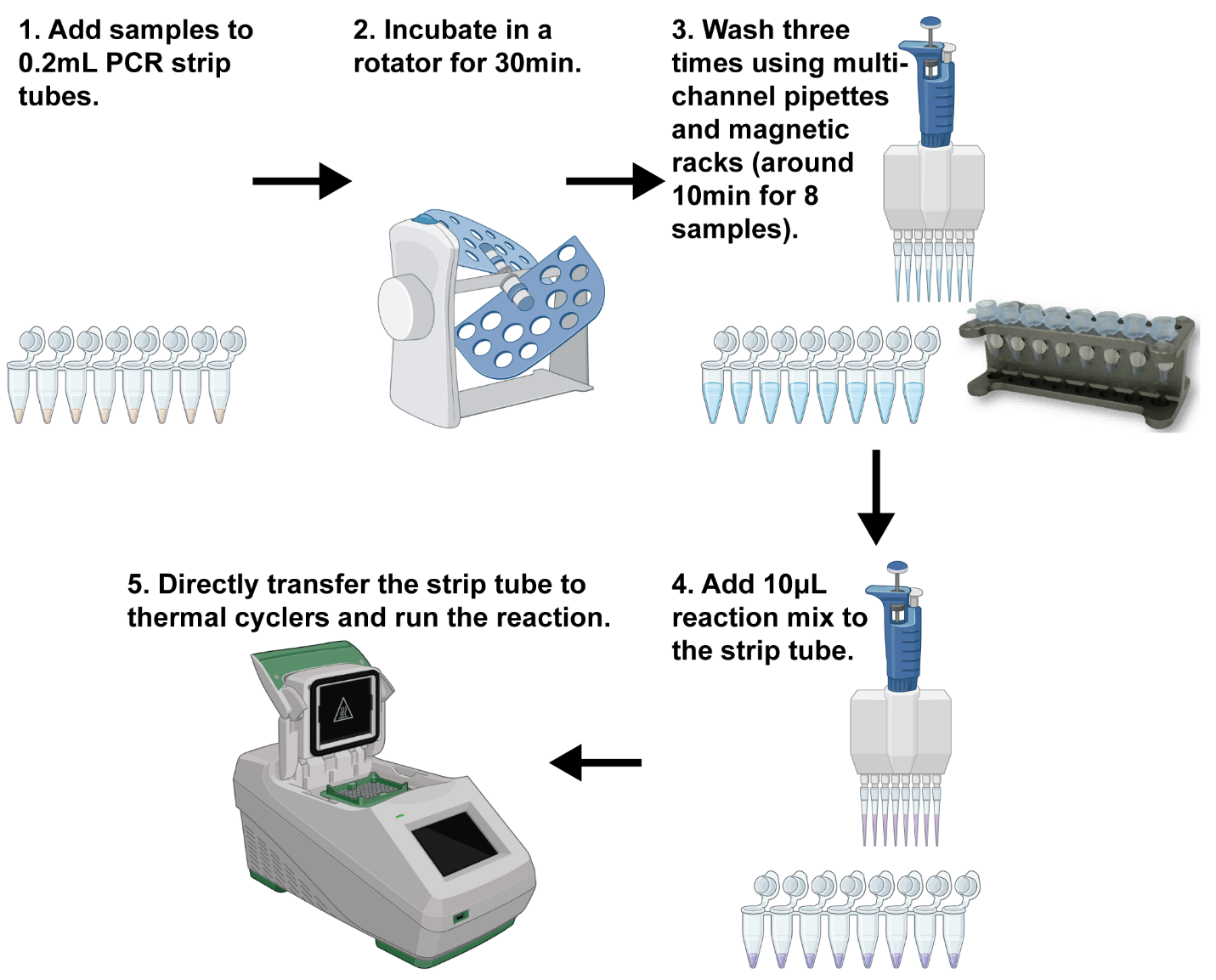

Figure S9 High-throughput testing with the integrated workflow by using strip tubes and multi-channel pipettes.

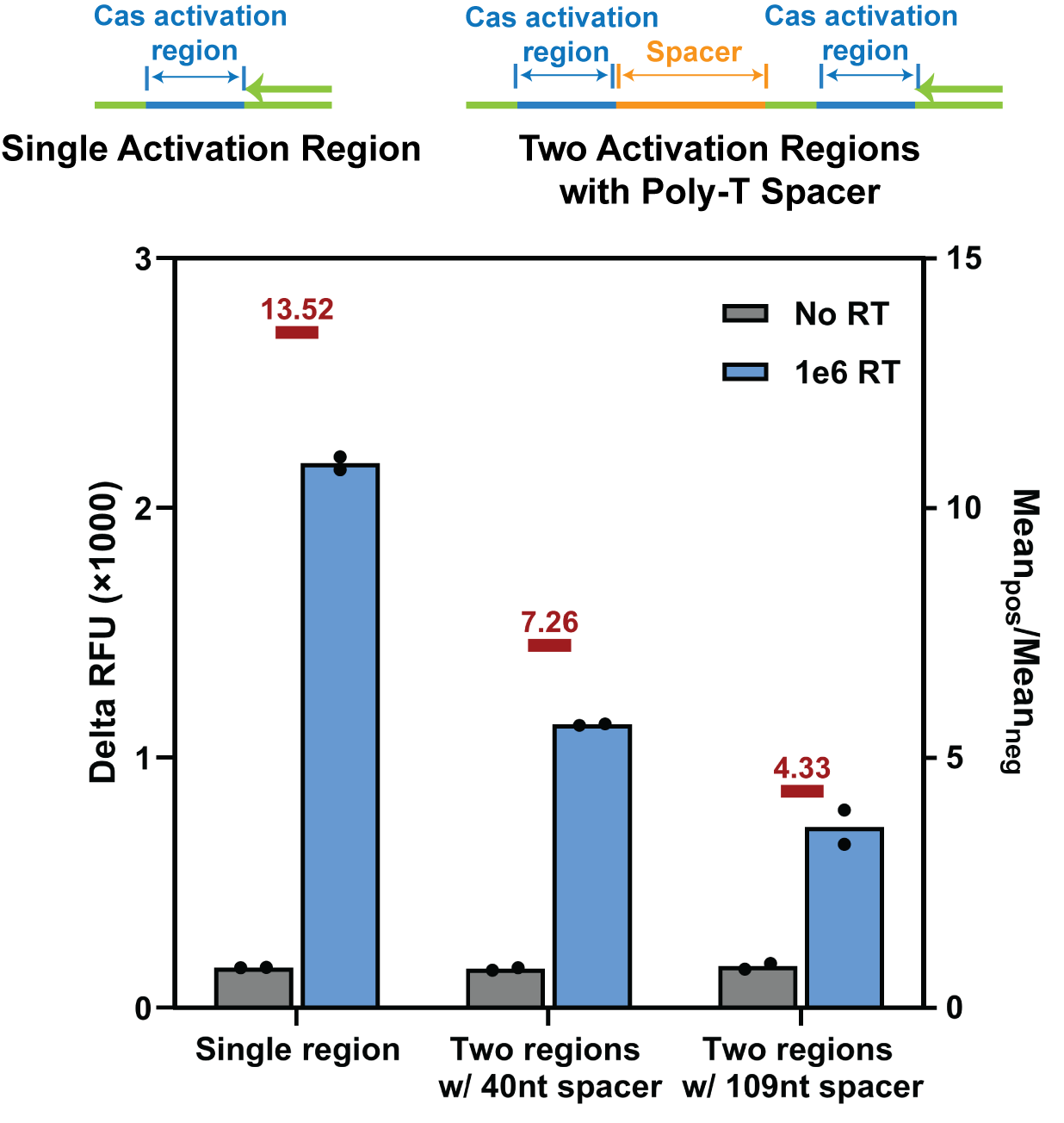

Figure S10 Delta RFU during 2hr incubation in CasPART of using substrates with two activation regions versus a single activation region. A poly-T spacer was included between activation regions to avoid steric hindrance.

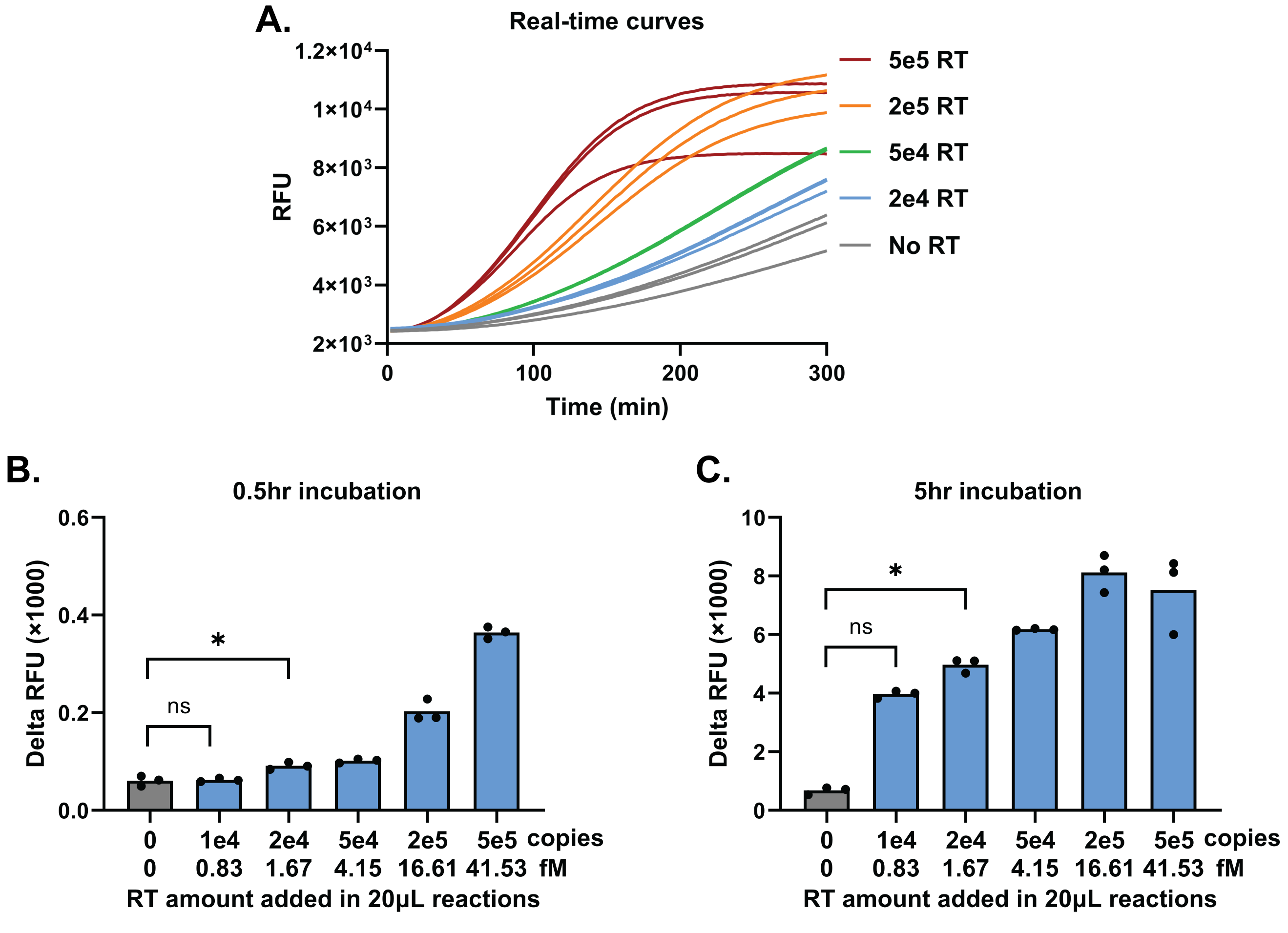

Figure S11 (A) Real-times curves of final CasPART performance with HIV RT enzyme in buffers during 5hr incubation. (B) Delta RFU during 0.5hr incubation. (C) Delta RFU during 5hr incubation. The LoD maintained 2e4 copies of HIV RT enzyme in both cases. Statistical significance was determined by unpaired t tests with Welch’s correction and conducted in GraphPad Prism 10. * indicates p < 0.05.

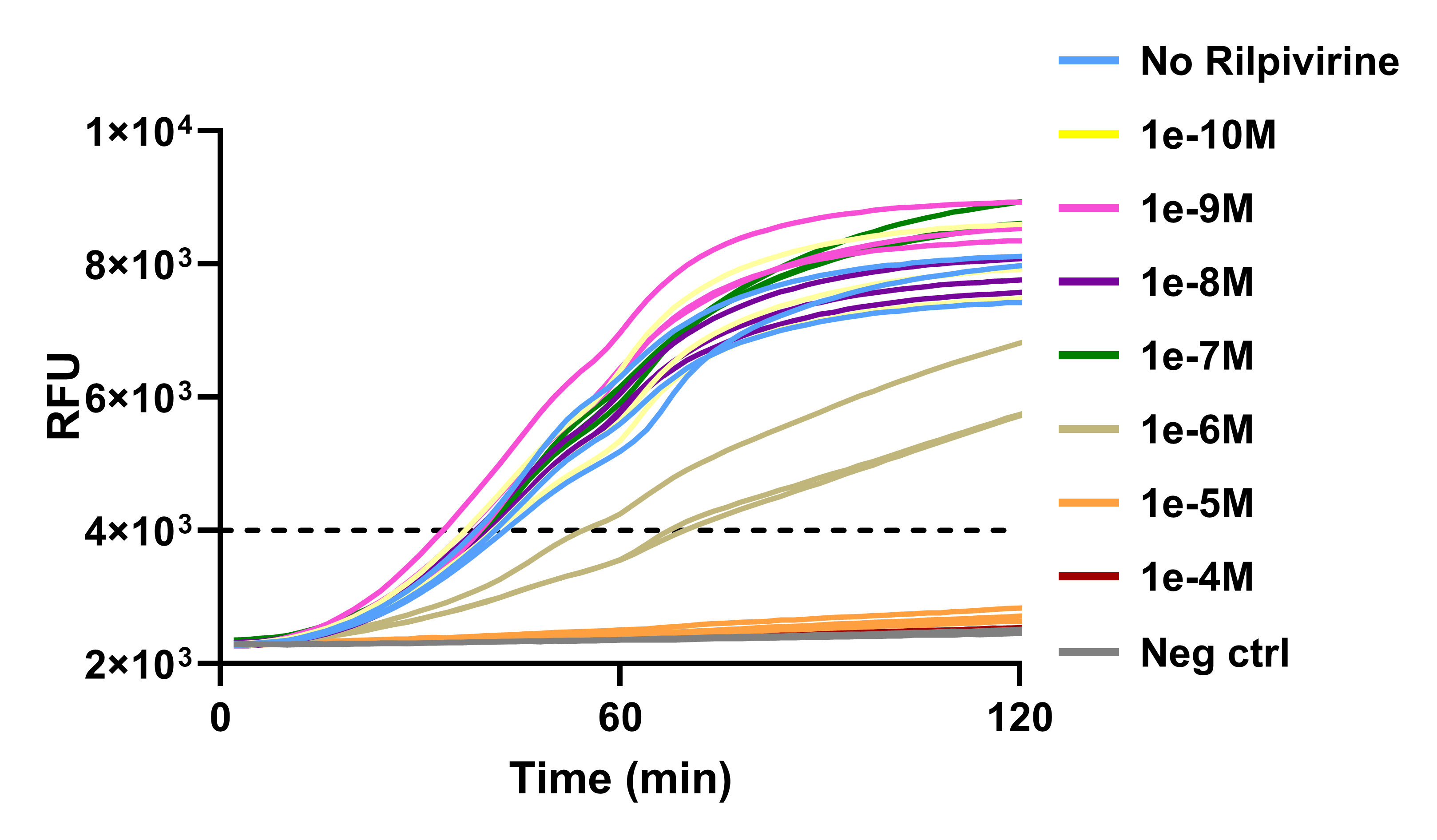

Figure S12 CasPART real-time curves of HIV RT enzyme with different concentrations of rilpivirine. The negative controls included no HIV RT enzyme or rilpivirine, while all other samples included 1e7 copies of HIV RT enzyme.

Supplementary Table S1 The fitted rates of HIV RT enzyme with different concentrations of rilpivirine.

| Rilpivirine conc. (M) | Replicate #1 | | Replicate #2 | | Replicate #3 | |
| --- | --- | --- | --- | --- | --- | --- |
|  | **Fitted rate** | **R squared** | **Fitted rate** | **R squared** | **Fitted rate** | **R squared** |
| 0 | 1.453 | 0.9995 | 1.272 | 0.9997 | 1.155 | 0.9998 |
| 1e-10 | 1.585 | 0.9997 | 1.221 | 0.9999 | 1.283 | 0.9997 |
| 1e-9 | 1.909 | 0.9998 | 1.563 | 0.9999 | 1.440 | 0.9988 |
| 1e-8 | 1.406 | 0.9999 | 1.322 | 0.9997 | 1.291 | 0.9995 |
| 1e-7 | 1.263 | 0.9996 | 1.271 | 0.9999 | 1.294 | 0.9998 |
| 1e-6 | 0.2690 | 0.9991 | 0.3021 | 0.9982 | 0.6275 | 0.9990 |
| 1e-5 | 0.01818 | 0.9980 | 0.01413 | 0.9973 | 0.01545 | 0.9966 |
| 1e-4 | 0.01082 | 0.9980 | 0.01017 | 0.9986 | 0.009490 | 0.9984 |
| Neg ctrl  (no RT, no rilpivirine) | 0.006392 | 0.9966 | 0.008912 | 0.9973 | 0.005319 | 0.9979 |

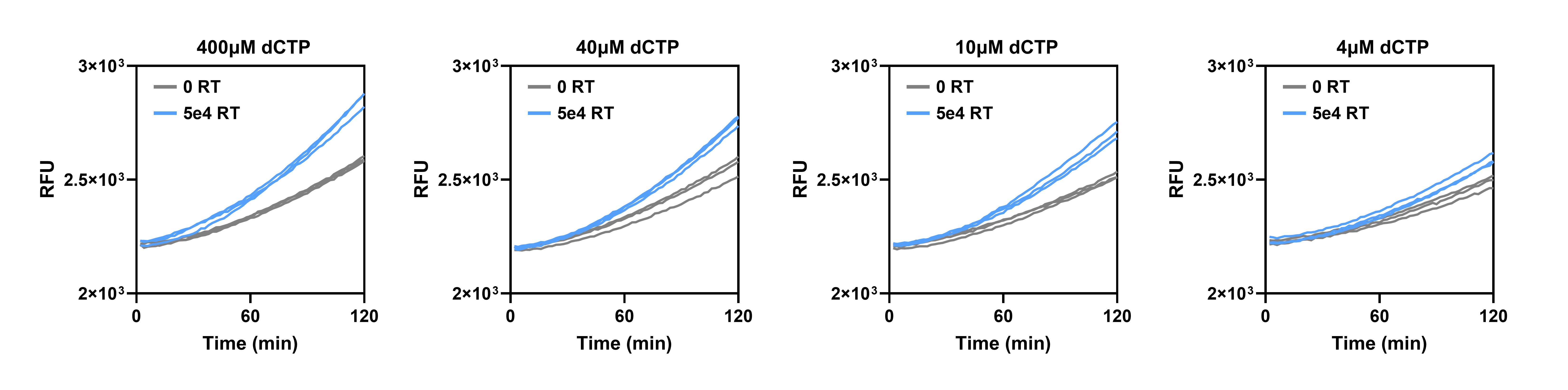

Figure S13 CasPART real-time curves of negative controls and 5e4 RT samples with different concentrations of dCTP used in the assay. The concentrations of dATP, dGTP, and dTTP were kept constant at 400μM.

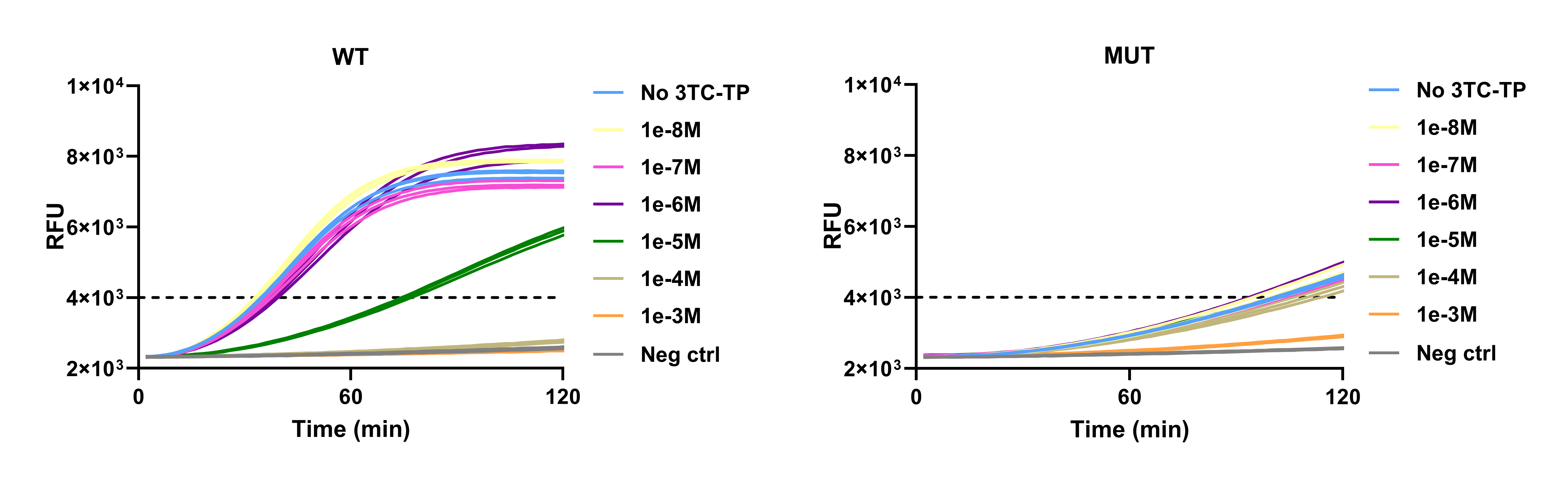

Figure S14 CasPART real-time curves of wild-type (WT) or mutant (MUT) HIV RT enzyme with different concentrations of 3TC-TP. The negative controls included no HIV RT enzyme or 3TC-TP, while the HIV RT enzyme copy number in other samples was set at 1e7.

Supplementary Table S2 The fitted rates of WT HIV RT enzyme with different concentrations of 3TC-TP.

| 3TC-TP conc. (M) | Replicate #1 | | Replicate #2 | | Replicate #3 | |
| --- | --- | --- | --- | --- | --- | --- |
|  | **Fitted rate** | **R squared** | **Fitted rate** | **R squared** | **Fitted rate** | **R squared** |
| 0 | 1.601 | 1.000 | 1.587 | 1.000 | 1.644 | 1.000 |
| 1e-8 | 1.833 | 1.000 | 1.734 | 1.000 | 1.798 | 1.000 |
| 1e-7 | 1.388 | 0.9999 | 1.543 | 1.000 | 1.470 | 1.000 |
| 1e-6 | 1.234 | 1.000 | 1.300 | 0.9999 | 1.405 | 1.000 |
| 1e-5 | 0.2853 | 0.9998 | 0.3053 | 0.9998 | 0.3044 | 0.9998 |
| 1e-4 | 0.02427 | 0.9997 | 0.02674 | 0.9997 | 0.02535 | 0.9997 |
| 1e-3 | 0.005489 | 0.9987 | 0.006874 | 0.9980 | 0.006776 | 0.9983 |
| Neg ctrl  (no RT, no 3TC-TP) | 0.01139 | 0.9991 | 0.01109 | 0.9992 | 0.009856 | 0.9989 |

Supplementary Table S3 The fitted rates of MUT HIV RT enzyme with different concentrations of 3TC-TP.

| 3TC-TP conc. (M) | Replicate #1 | | Replicate #2 | | Replicate #3 | |
| --- | --- | --- | --- | --- | --- | --- |
|  | **Fitted rate** | **R squared** | **Fitted rate** | **R squared** | **Fitted rate** | **R squared** |
| 0 | 0.1551 | 0.9996 | 0.16 | 0.9999 | 0.1628 | 0.9999 |
| 1e-8 | 0.1573 | 0.9999 | 0.1679 | 0.9999 | 0.1846 | 0.9999 |
| 1e-7 | 0.1488 | 0.9999 | 0.1593 | 0.9999 | 0.1635 | 0.9999 |
| 1e-6 | 0.1857 | 1.000 | 0.1837 | 0.9999 | 0.1889 | 0.9998 |
| 1e-5 | 0.1577 | 0.9995 | 0.1573 | 0.9999 | 0.1596 | 0.9999 |
| 1e-4 | 0.1248 | 0.9999 | 0.136 | 0.9999 | 0.1413 | 0.9999 |
| 1e-3 | 0.03316 | 0.9998 | 0.03598 | 0.9998 | 0.03657 | 0.9996 |
| Neg ctrl  (no RT, no 3TC-TP) | 0.007029 | 0.9982 | 0.01027 | 0.9991 | 0.01024 | 0.9978 |

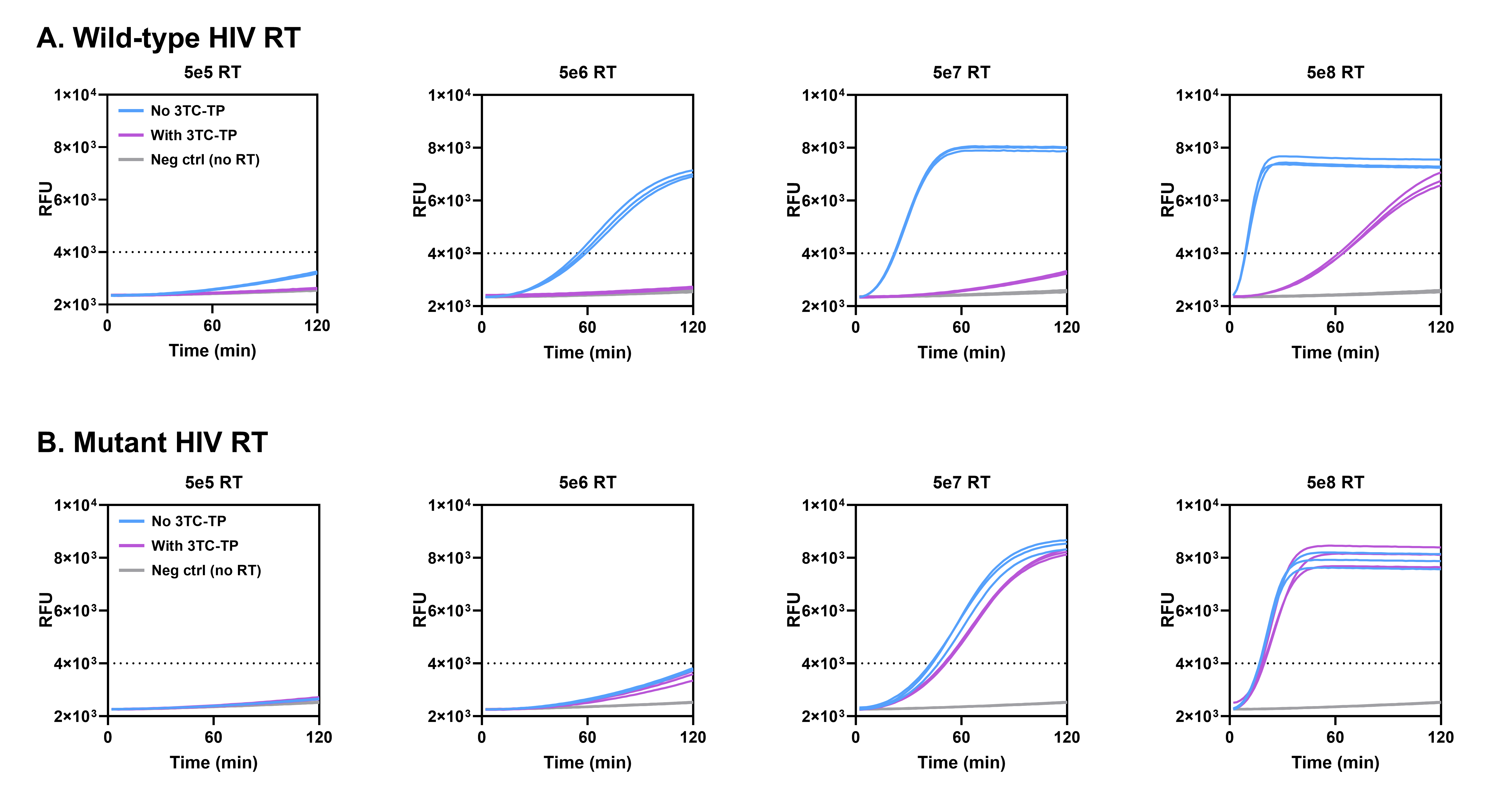

Figure S15 CasPART real-time curves of wild-type or mutant HIV RT enzyme at different copy numbers with or without 10^-4^ M 3TC-TP.

Supplementary Table S4 The fitted rates of WT HIV RT enzyme without 3TC-TP.

| RT copy number | Replicate #1 | | Replicate #2 | | Replicate #3 | |
| --- | --- | --- | --- | --- | --- | --- |
|  | **Fitted rate** | **R squared** | **Fitted rate** | **R squared** | **Fitted rate** | **R squared** |
| 0 | 0.008661 | 0.9984 | 0.008677 | 0.9979 | 0.008136 | 0.9980 |
| 5e5 | 0.05549 | 0.9997 | 0.06281 | 0.9998 | 0.06178 | 0.9998 |
| 5e6 | 0.6824 | 0.9998 | 0.6105 | 0.9998 | 0.5751 | 0.9998 |
| 5e7 | 4.155 | 0.9998 | 4.238 | 0.9998 | 4.369 | 0.9998 |
| 5e8 | 26.43 | 1.000 | 24.72 | 1.000 | 27.19 | 0.9999 |

Supplementary Table S5 The fitted rates of WT HIV RT enzyme with 3TC-TP.

| RT copy number | Replicate #1 | | Replicate #2 | | Replicate #3 | |
| --- | --- | --- | --- | --- | --- | --- |
|  | **Fitted rate** | **R squared** | **Fitted rate** | **R squared** | **Fitted rate** | **R squared** |
| 0 | 0.01135 | 0.9982 | 0.01242 | 0.9986 | 0.01089 | 0.9986 |
| 5e5 | 0.01301 | 0.9991 | 0.01284 | 0.9991 | 0.01249 | 0.9987 |
| 5e6 | 0.01678 | 0.9992 | 0.01717 | 0.9993 | 0.0184 | 0.9992 |
| 5e7 | 0.06716 | 0.9999 | 0.06163 | 0.9999 | 0.06022 | 0.999 |
| 5e8 | 0.4932 | 0.9998 | 0.4579 | 0.9999 | 0.4965 | 0.9999 |

Supplementary Table S6 The fitted rates of MUT HIV RT enzyme without 3TC-TP.

| RT copy number | Replicate #1 | | Replicate #2 | | Replicate #3 | |
| --- | --- | --- | --- | --- | --- | --- |
|  | **Fitted rate** | **R squared** | **Fitted rate** | **R squared** | **Fitted rate** | **R squared** |
| 0 | 0.0107 | 0.9987 | 0.009737 | 0.9989 | 0.009624 | 0.9981 |
| 5e5 | 0.02088 | 0.9996 | 0.01863 | 0.9998 | 0.02045 | 0.9995 |
| 5e6 | 0.1097 | 0.9999 | 0.1058 | 1.000 | 0.1135 | 0.9999 |
| 5e7 | 1.119 | 0.9996 | 1.082 | 0.9999 | 0.9722 | 0.9995 |
| 5e8 | 7.264 | 0.9998 | 7.484 | 0.9999 | 7.168 | 0.9998 |

Supplementary Table S7 The fitted rates of MUT HIV RT enzyme with 3TC-TP.

| RT copy number | Replicate #1 | | Replicate #2 | | Replicate #3 | |
| --- | --- | --- | --- | --- | --- | --- |
|  | **Fitted rate** | **R squared** | **Fitted rate** | **R squared** | **Fitted rate** | **R squared** |
| 0 | 0.01284 | 0.9988 | 0.00996 | 0.9990 | 0.01318 | 0.9989 |
| 5e5 | 0.02402 | 0.9996 | 0.02016 | 0.9994 | 0.02406 | 0.9997 |
| 5e6 | 0.1048 | 0.9999 | 0.08065 | 0.9999 | 0.0976 | 1.000 |
| 5e7 | 0.8067 | 0.9996 | 0.8133 | 0.9997 | 0.8147 | 0.9997 |
| 5e8 | 5.435 | 0.9999 | 6.670 | 0.9998 | 5.364 | 0.9999 |

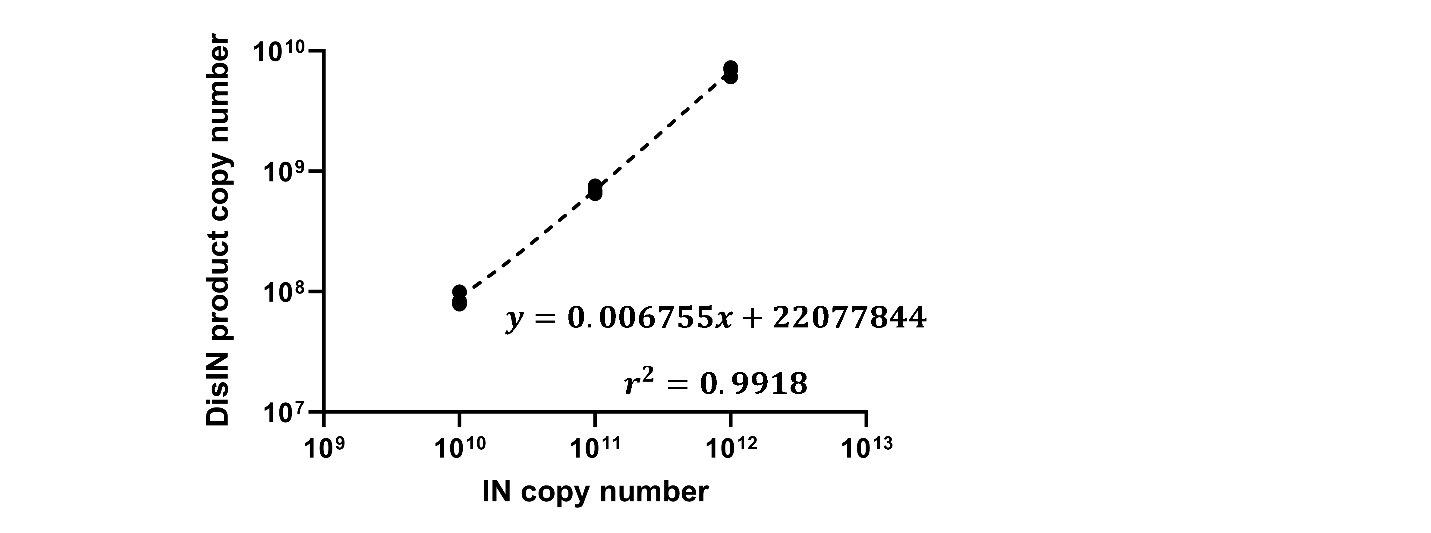

Figure S16 The linear correlation of disintegration product copy number versus integrase copy number, as quantified by qPCR.

**Sequence Table**

| **Characterization of cDNA generation by HIV RT enzyme with qPCR** | | |
| --- | --- | --- |
| Sequence #1 | RNA template  (all RNA bases) | GGGAGACAUUAGCCAUUUCAACCCAUGCGUUUGAGGAGAAGCGCUUUCUGAUAACCGGUGGUCUCCCAUCAGGUUGUGCAGCGACCUCAAUGCUAAACACUAUAAUGAAUAAUAUAAUAAUUAGGGCGGGUUUGUAUCUCACGUAUAAAAAUUUUGAAUUUGAUGAUGUGAAGGUGUUGUCGUACGGAGAUGAUCUCCUUGUGGCCACAAAUUACCAAUUGGAUUUUGAUAAGGUGAGAGCAAGCCUCGCAAAGACAGGAUAUAAGAUAACUCCCGCUAACACAACUUCUACCUUUCCUCUUAAUUCGACGCUUGAAGACGUUGUCUUCUUAAAAAGAAAGUUUAAGAAAGAGGGCCCUCUGUAUCGGCCUGUCAUGAAC |
|  | RT primer/  PCR Rev primer | CGTCGAATTAAGAGGAAAGGTAG |
|  | PCR Fwd primer | TTTGATAAGGTGAGAGCAAGC |
| Sequence#2 | RNA template  (all RNA bases) | GCCGGCCUAGCUGUGAUAAGUCUAUCUCUCUUUCCACGGGCUAUGGAAUCGCUACAGGAGGUUAGAGAGUCGAUACGUGCGCUAUGUGCUCGGCUAUAUCUAAGCAAGCUUUCCUCCGAUACCCAUCGCGUAGAGGA |
|  | RT primer/  PCR Rev primer | GCTCACACTACTCCTCTACGC |
|  | PCR Fwd primer | CCGGCCTAGCTGTGATAAGTC |
| Sequence #3 | RNA template  (all RNA bases) | GUGGGAAGUUCAAUUAGGAAUACGAUGCAUAUUUUUCAGUUCCCUUAGAUCUGCAUUCACCAUACCUAGUAAACAAUGAGACACCAGGAGAUAUCAGUACAAUGUGCUGAUGGAAAGGAUCACCAGCAAUAUUAGCAUGACAAA |
|  | RT primer/  PCR Rev primer | TGGGAAGTTCARTTAGGAATAC |
|  | PCR Fwd primer | GGCTCTAAGATTTTTGTCATGCT |
| Sequence #4 | RNA template  (all RNA bases) | GUGUUGUUCUGUCUGUCUUCCCACCUCCUUCUCCUUCUUCUUGAGUGAAGUGUGUGAGAGGUGGUGAGAGUGAGCACUUCCUCUUCUUCUGUUGUUGUCUCCCUCUUCUUCGUGGUGAGUG |
|  | RT primer/  PCR Rev primer | CACTCACTCACACTCACCAC |
|  | PCR Fwd primer | GTGTTGTTCTGTCTGTCTTC |
| **LamPART with real-time fluorescence detection** | | |
| Sequence #2, full length  (all RNA bases) | | GCCGGCCUAGCUGUGAUAAGUCUAUCUCUCUUUCCACGGGCUAUGGAAUCGCUACAGGAGGUUAGAGAGUCGAUACGUGCGCUAUGUGCUCGGCUAUAUCUAAGCAAGCUUUCCUCCGAUACCCAUCGCGUAGAGGAGUAGUGUGAGCGUGGGAUAUGUACGACGACCG |
| Sequence #2, truncated design  (all RNA bases) | | GCCGGCCUAGCUGUGAUAAGUCUAUCUCUCUUUCCACGGGCUAUGGAAUCGCUACAGGAGGUUAGAGAGUCGAUACGUGCGCUAUGUGCUCGGCUAUAUCUAAGCAAGCUUUCCUCCGAUACCCAUCGCGUAGAGGA |
| LAMP primers | FIP | GTGCTCGGCTATATCTAAGCAGCTCACACTACTCCTCTACGC |
|  | BIP | ATAGCGCACGTATCGACTCTCTATCTCTCTTTCCACGGGCTA |
|  | UDPtag-LF | ACCACACCTACCACCACTAATAACTAAAGCTTTCCTCCGATACCCATC |
|  | LB | TAACCTCCTGTAGCGATTCCA |
|  | B3 | CCGGCCTAGCTGTGATAAGTC |
|  | F3  (not included in final LamPART) | CGGTCGTCGTACATATCCCAC |
| Probes | UDP quencher | CTGTCGTAGGTGGTTGGGTTCGGAAGTGGTCAGG/3IABkFQ/ |
|  | UDP fluorophore | /56-FAM/CCTGACCACTTCCGAACCCAACCACCTACGACAGACCACACCTACC  ACCACTAATAACTAA |
| Aptamer | TQ21 | TTCTCGGTTGGTCTCTGGCGGAGCGATCATCTCAGAGCATTCTTAGCGTTTTGTTCTTGTGTATGATTCGCTTTTCCC |
| Dockers | LB docker | GCTACAGGAGGTTA/3ddC/ |
|  | B3 docker | ACAGCTAGGCCGG/3ddC/ |
| **LamPART with endpoint lateral flow detection** | | |
| FIP | | /5BiosG/GTGCTCGGCTATATCTAAGCAGCTCACACTACTCCTCTACGC |
| BIP | | /56-FAM/ATAGCGCACGTATCGACTCTCTATCTCTCTTTCCACGGGCTA |

| **CasPART** | |
| --- | --- |
| **Substrate preference** | |
| Primer | CGTCGAATTAAGAGGAAAGGTAG |
| crRNA-1  (all RNA bases) | /AlTR1/UAAUUUCUACUAAGUGUAGAUAUGAUGUGAAGGUGUUGUCG/AlTR2/ |
| DNA substrate | TTTTTTTTTTTTTGATGATGTGAAGGTGTTGTCGCTACCTTTCCTCTTAATTCGACG |
| Chimeric substrate  (bases with prefix ‘r’ are RNA bases) | rUrUrUrUrUrUrUrUrUrUTTTGATGATGTGAAGGTGTTGTCGrCrUrArCrCrUrUrUrCrCrUrCrUrUrArArUrUrCrGrArCrG |
| RNA substrate: sequence #1 used in characterization of cDNA generation by HIV RT enzyme with qPCR  (all RNA bases) | GGGAGACAUUAGCCAUUUCAACCCAUGCGUUUGAGGAGAAGCGCUUUCUGAUAACCGGUGGUCUCCCAUCAGGUUGUGCAGCGACCUCAAUGCUAAACACUAUAAUGAAUAAUAUAAUAAUUAGGGCGGGUUUGUAUCUCACGUAUAAAAAUUUUGAAUUUGAUGAUGUGAAGGUGUUGUCGUACGGAGAUGAUCUCCUUGUGGCCACAAAUUACCAAUUGGAUUUUGAUAAGGUGAGAGCAAGCCUCGCAAAGACAGGAUAUAAGAUAACUCCCGCUAACACAACUUCUACCUUUCCUCUUAAUUCGACGCUUGAAGACGUUGUCUUCUUAAAAAGAAAGUUUAAGAAAGAGGGCCCUCUGUAUCGGCCUGUCAUGAAC |
| **Poly-T tail** | |
| crRNA-1  (all RNA bases) | /AlTR1/UAAUUUCUACUAAGUGUAGAUAUGAUGUGAAGGUGUUGUCG/AlTR2/ |
| dsDNA substrate  with no tail | Strand #1:  TTTGATGATGTGAAGGTGTTGTCGCTACCTTTCCTCTTAATTCGACG  Strand #2:  AAACTACTACACTTCCACAACAGCGATGGAAAGGAGAATTAAGCTGC |
| dsDNA substrate  with 10-T tail | Strand #1: TTTTTTTTTTTTTGATGATGTGAAGGTGTTGTCGCTACCTTTCCTCTTAATTCGACG  Strand #2:  AAAAAAAAAAAAACTACTACACTTCCACAACAGCGATGGAAAGGAGAATTAAGCTGC |
| dsDNA substrate  with 20-T tail | Strand #1: TTTTTTTTTTTTTTTTTTTTTTTGATGATGTGAAGGTGTTGTCGCTACCTTTCCTCTTAATTCGACG  Strand #2:  AAAAAAAAAAAAAAAAAAAAAAACTACTACACTTCCACAACAGCGATGGAAAGGAGAATTAAGCTGC |
| **Primer length** | |
| Substrate | TTTTTTTTTTTTTGAGGAGGTTAGAGAGTCGATACGTGCGCTATGTGCTCGGCT |
| crRNA-2  (all RNA bases) | /AlTR1/UAAUUUCUACUAAGUGUAGAUAGGAGGUUAGAGAGUCGAUA/AlTR2/ |
| 20nt primer | AGCCGAGCACATAGCGCACG |
| 20+5nt primer | AGCCGAGCACATAGCGCACGTATCG |
| 20+10nt primer | AGCCGAGCACATAGCGCACGTATCGACTCT |
| 20+15nt primer | AGCCGAGCACATAGCGCACGTATCGACTCTCTAAC |
| **Final assay** | |
| Substrate | TTTTTTTTTTTTTGAGGAGGTTAGAGAGTCGATACGTGCGCTATGTGCTCGGCT |
| Primer | AGCCGAGCACATAGCGCACGTATCGACTCT |
| crRNA-2  (all RNA bases) | /AlTR1/UAAUUUCUACUAAGUGUAGAUAGGAGGUUAGAGAGUCGAUA/AlTR2/ |
| Reporter | /56-FAM/TTATT/3IABkFQ/ |
| **HIV integrase detection** | |
| NTS | TCTCTCTCTCTTTGAGGAGGTTAGAGAGTCGATACGTGCGCTAT |
| TS-3-Tail | ACTGCTAGAGATTTTCCACA |
| TS-3 (10nt distance; final) | TGTGGAAAATCTCTAGCACTAACCTCCTCAAAGAGAGAGAGA |
| TS-5 (10nt distance; final) | ATAGCGCACGTATCGACTCT |
| TS-3 (0nt distance) | TGTGGAAAATCTCTAGCACAAAGAGAGAGAGA |
| TS-5 (0nt distance) | ATAGCGCACGTATCGACTCTCTAACCTCCT |
| TS-3 (5nt distance) | TGTGGAAAATCTCTAGCACTCCTCAAAGAGAGAGAGA |
| TS-5 (5nt distance) | ATAGCGCACGTATCGACTCTCTAAC |
| TS-3 (15nt distance) | TGTGGAAAATCTCTAGCAACTCTCTAACCTCCTCAAAGAGAGAGAGA |
| TS-5 (15nt distance) | ATAGCGCACGTATCG |
